# Persistence of targetable lesions, predicted therapy sensitivity and proteomes through disease evolution in pediatric acute lymphoblastic leukemia

**DOI:** 10.1101/2022.03.04.22271927

**Authors:** Amanda C. Lorentzian, Jenna Rever, Enes K. Ergin, Meiyun Guo, Neha M. Akella, Nina Rolf, C. James Lim, Gregor S.D. Reid, Christopher A. Maxwell, Philipp F. Lange

## Abstract

Childhood acute lymphoblastic leukemia (ALL) genomes show that relapses often arise from subclonal outgrowths. However, the impact of clonal evolution on the actionable proteome and response to targeted therapy is not known. Here, we present a comprehensive retrospective analysis of paired ALL diagnosis and relapsed specimen. Targeted next generation sequencing and proteome analysis indicated persistence of actionable genome variants and stable proteomes through disease progression. Paired viably-frozen biopsies showed high correlation of drug response to variant-targeted therapies but *in vitro* selectivity was low. Proteome analysis prioritized PARP1 as a new pan-ALL target candidate needed for survival following cellular stress; diagnostic and relapsed ALL samples demonstrated robust sensitivity to treatment with two PARP1/2 inhibitors. Together, these findings support initiating prospective precision oncology approaches at ALL diagnosis and emphasize the need to incorporate proteome analysis to prospectively determine tumor sensitivities, which are likely to be retained at disease relapse.

**STATEMENT OF SIGNIFICANCE:** We discover that disease progression and evolution in pediatric acute lymphoblastic leukemia is defined by the persistence of targetable genomic variants and stable proteomes, which reveal pan-ALL target candidates. Thus, personalized treatment options in childhood ALL may be improved with the incorporation of prospective proteogenomic approaches initiated at disease diagnosis.

## INTRODUCTION

Relapsed cancer is a leading disease-related cause of death for children and adolescents (1). Targeting the specific molecular changes that arise in cancer cells may improve patient survival (2); for this reason, clinical trials centred on next generation sequencing (NGS)-based identification of biomarkers and targetable pathways are currently establishing patient enrolment strategies, clinical protocols, and critical safety data for personalized therapies. Most trials are currently limited to high-risk or recurrent disease. However, fast progressing disease often limits successful treatment options. In certain cases, precision oncology trials may be initiated at diagnosis. But, a major challenge for prospective precision oncology approaches is our limited understanding of the persistence or evolution of targetable lesions and their associated proteins or pathways, and responses to targeted agents, that may be gained or lost at relapse.

There is now a wealth of publicly available data for genomic characterization of paired diagnosis and relapse specimens from children with cancer. Clonal evolution does occur in pediatric leukemia wherein a minor clone, present only at low frequencies at diagnosis, is selected at time of relapse (3–5). While these studies show evolution in single nucleotide variants, often through chemotherapy-induced mutation (6), structural variants change less frequently with leukemia progression.

Few studies have yet investigated how protein levels change from diagnosis to relapse, especially pertaining to therapeutic targets. A previous proteomic analysis of matched diagnostic and relapsed B-cell precursor acute lymphoblastic leukemia (ALL) specimens, sourced from pediatric and adult patients, observed increased protein levels in specific pathways at relapse, including glycoloysis, phosphate pentose pathway and metabolic pathways that may contribute to chemo-resistance, but it was not specific to pediatric ALL and was limited to approximately 1400 proteins (7). Since proteins are the actual therapeutic targets (8,9), it is crucial to better understand the pediatric tumor proteome to determine how the response to therapy may change through progression. Here, we present a comprehensive interrogation of ALL proteogenome dynamics from diagnosis to relapse in paired patient specimens, specifically to understand how cancer-driving and potentially targetable lesions persist or differentiate through disease progression.

## RESULTS

### Next-generation sequencing (NGS) reveals stability of affected genes through ALL disease progression

To examine proteogenomic evolution in relapsed pediatric ALL, we sourced 71 bone marrow biopsies (n=44 ALL at initial diagnosis (Dx), n=21 ALL at relapse (R), n=6 non-cancer bone marrow) from pediatric patients seen at BC Children’s Hospital (BCCH) (Supplementary Table S1) and publicly available whole-exome sequencing (WES) data from 138 specimens collected and analyzed by the St. Jude’s Children’s Research Hospital (SJH) (Fig. 1A).

**Figure 1:**
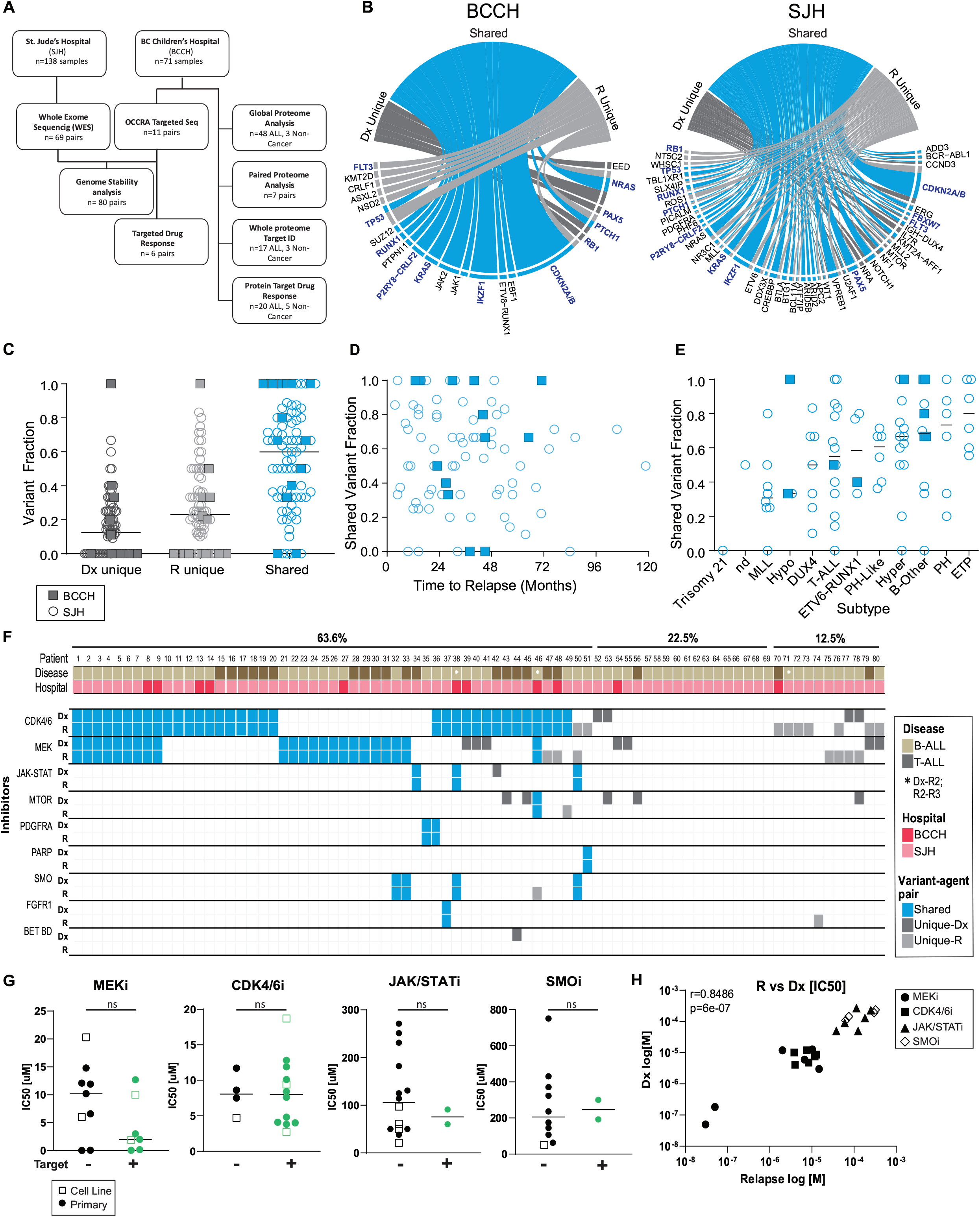
Stability of affected genes and targeted drug response through ALL disease progression. **A**. Flow-chart depicting the number of samples from each cohort that were included for each analysis. **B**. Circos plot for all mutations identified in the BCCH and SJH cohorts demonstrating mutations that were Dx unique (dark grey), R unique (light grey), or shared between Dx and R samples (blue). Genes in blue text indicate genes with detected lesions in both cohorts. **C**. Fraction of variants identified as D unique, R1 unique, or shared within paired samples sourced from 80 ALL patients (n=11 from BCCH represented by squares, n=69 from SJH represented by circles). The black bar represents the median of the population. **D**. Dot plot for fraction of shared variants versus time to relapse for 80 ALL patients (n= 11 from BCCH represented by squares, n=69 from SJH represented by circles). **E**. Dot plot for fraction of shared variants separated by disease subtype for 80 ALL patients (n=11 from BCCH represented by squares, n=69 from SJH represented by circles). The black bar represents the median of the population. **F**. Predicted sensitivity to targeted agents in paired Dx-R samples (or Dx-R2, R2-R3 indicated by an asterisk) taken from 80 ALL patients treated at BCCH (n=11, red) or SJH (n=69, dark pink). B-ALL (light brown) and T-ALL (dark brown) samples are indicated. Shared variants (blue), Dx unique variants (dark grey), or R unique variants (light grey) are indicated. Agent-variant pairs were assigned following the strategy outlined in the Pediatric Match Trial (9). **G**. Measured IC50 values for viably frozen ALL samples from patients treated at BCCH (n=6 patients) and representative cell lines. ALL samples were co-cultured with hTERT-MSCs and separately treated with four inhibitors, MEKi, CDK4/6i, JAK/STATi, and SMOi. IC50 [µM] are colored by most sensitive in yellow to least sensitive in blue. Measurements represent the mean of n=2 replica wells from a single experiment. **H**. Dot plots for IC50 [µM] values measured for primary samples or cell lines for each drug. Samples are separated based on the presence (green) or absence (black) of a genomic variant predicted to augment drug sensitivity (ns= not significant by unpaired t-test). **I**. Correlation of IC50 [M] values measured for paired Dx and R paired samples. Individual drugs are indicated by unique identifiers. r= Pearson correlation coefficient.

At time of relapse of pediatric ALL, clonal evolution is frequently convergent, and includes the outgrowth of clones defined by a different mutational site within the same affected gene; for example, a relapse clone with KRAS.A146T replaces the diagnostic KRAS.G12D clone (3). Since alternate pathogenic mutations in the same gene often serve as biomarkers for the same targeted treatment, we focused our analysis on the affected gene, rather than the mutation site. We first explored the mutational landscape in paired progression samples (n=10 B-ALL, n=1 T-ALL) via targeted, pediatric cancer-focused NGS analysis (10). Across the BCCH cohort, we detected recurrent copy number variants (CNV) or single nucleotide variants (SNV) in *CDKN2A/B, NRAS, KRAS, IKZF, JAK1*, and *JAK2* (Supplementary Fig. S1, Supplementary Figure S2, and Supplementary Table S2), which are commonly mutated in pediatric ALL samples (9). Nine of the eleven patients had at least 50% retention of mutations and four of these patients had 100% retention of mutations (Supplementary Fig. S1).

We evaluated the lesions grouped by detection only at diagnosis (Dx unique), only at relapse (R unique), or at both timepoints (shared). Here we found 67% (30 of 45) of affected genes were shared between paired diagnosis and relapse samples in the BCCH cohort (Fig. 1B). To determine the generalizability of this finding, we mined public NGS datasets from ALL cases (n=49 B-ALL; n=20 T-ALL) treated at St. Jude’s Hospital (SJH) (Supplementary Table S3) (11). Samples collected from BCCH and SJH showed similar distributions of affected genes that were shared between time-points or were unique to Dx or R (Fig. 1B and Supplementary Table S4), with the majority of variants shared between paired diagnosis and relapse samples (Fig. 1C). In fact, the genes that were persistently mutated through disease progression were highly similar in both cohorts, including *CDKN2A/B, IKZF1*, and *N/KRAS*, with structural variants being retained with higher frequency than SNVs (Supplementary Fig. S4A-C). Surprisingly, retention of genetic lesions was not correlated with the time between diagnosis and first relapse, or between relapses (Fig. 1D and Supplementary Fig. S3D,E). However, we observed relatively higher lesion stability for Hyperdiploid (hyper), Philadelphia + (PH), and early T-cell precursor (ETP) sub-types (Fig. 1E).

### Matched patient-derived leukemic cells respond similarly to variant-selected agents

Persistence of affected genes suggests that sensitivity to precision therapies may also persist with disease progression. To examine this, we paired affected genes with targeted agents following the Pediatric MATCH strategy and evidence from clinical trials or case reports, as described (10). In the SJH and BCCH cohorts (n=80 paired samples), we found 64% of ALL patients (51 of 80) retained at least one variant-agent pairing at disease relapse; in fact, nearly 50% of patients (38 of 80) showed complete retention of variant-agent pairings through disease progression (Fig. 1F and Supplementary Table S5). Only five patients had variant-agent pairings unique to the diagnostic timepoint, while 13 patients harbored no targetable mutations (Fig. 1F).

We sourced viably-frozen bone marrow mononuclear cells (BM-MNC) from paired progression events for six patients treated at BCCH, including four paired BM-MNC that showed retention of all affected genes and two paired specimens with partial/no retention. Within this cohort, we identified four agents targeting affected genes (CDK4/6: Palbociclib; MEK: Trametinib, JAK/STAT: Ruxolitinib; SMO: Vismodegib) and then we treated viable patient specimens with graded doses of these agents. We used image-based drug screening of B-ALL cells co-cultured with mesenchymal stromal cells (MSC) to determine IC50 values for each sample after exposure (for 48-72 hours) to the four different targeted agents (Fig. 1G). Overall, the measured IC50 values revealed poor selectivity, with predicted responses to trametinib being an exception. Trametinib IC50 values were lower for patient samples predicted to be sensitive to MEK inhibition (Fig. 1G), although the difference was not significant (p=0.1322). In contrast, Palbociclib IC50 values did not differ within the primary ALL cohort (Fig. 1G) suggesting cytotoxicity is induced in a non-targeted manner by this agent. Overall, the measured IC50 values in relapsed samples correlated with values measured in matched diagnostic samples (Pearson’s r = 0.85, *p* = 6.0e-7, Fig. 1H). Therefore, our genomic analysis of pediatric ALL disease progression samples revealed stability of gene lesions that are known therapeutic targets. Drug sensitivities within matched diagnostic and relapsed samples were also highly correlated, though the drugs showed poor selectivity.

### Global proteome analysis shows stability through progression and groups cases with poor outcome

To determine whether the observed persistence of cancer-associated targetable genomic lesions and associated drug sensitivities is also characteristic of the proteome, we next conducted a comprehensive analysis of 48 primary (n=39 B-ALL, n=9 T-ALL) specimens from Dx and R, including 14 specimens from 6 patients with matched biopsies taken at diagnostic and subsequent relapse timepoints. Additionally, we included five pediatric ALL cell lines in our study (B-ALL=4, T-ALL=1). Our diverse cohort span the major cytogenetic groups and ages ranging from 2 years to 23 years. Male patients were moderately overrepresented (65% male compared to 35% female) (Fig. 2A, Supplementary Table S1).

**Figure 2:**
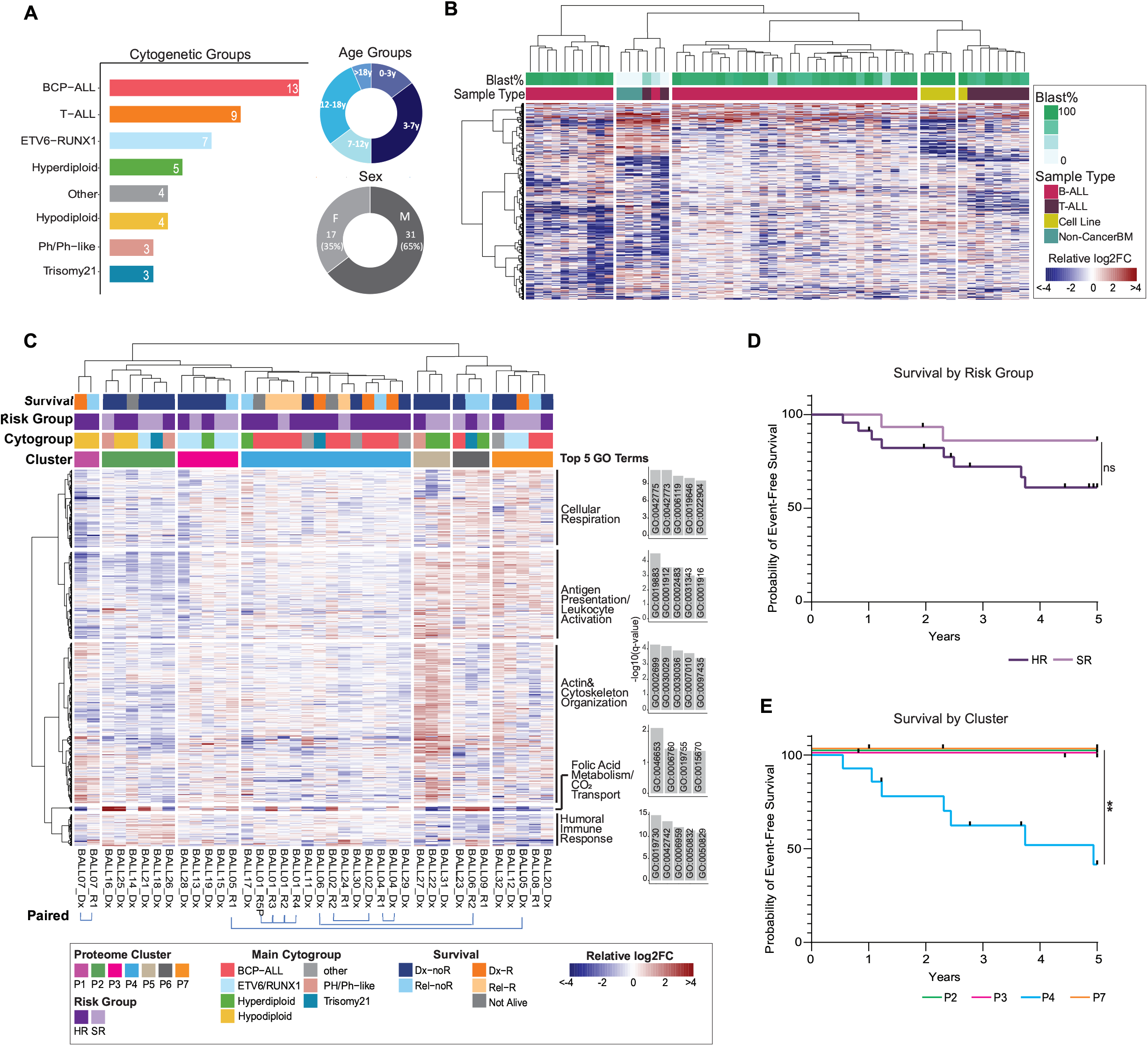
Global proteome analysis suggests shows stability through progression and groups cases with poor outcome. **A**. Descriptive summary of the cohort for proteome analysis. The bar plot represents the total number of each cytogenetic subtype and the donut plots represent the age (top) and sex (bottom) of the patients. **B**. Hierarchical clustering of 3907 variable proteins represented by the relative log2FC (protein intensity/median protein intensity). The color bars indicate sample type (bottom) and leukemic blast percentage (top). **C**. The major B-ALL clusters were selected for in-depth characterization. Hierarchical cluster of 935 proteins based on log2FC defined 7 sample clusters (horizontal) and 5 protein clusters (vertical). The remaining color bars indicate main cytogenetic subgroup (second from the bottom), followed by clinically assigned risk group (SR= standard risk, HR= high risk), and current survival status on top. The five most significant GO terms for each cluster of proteins were selected for visualizations. The annotation to the right of each cluster of proteins is the summary of the top significant terms. Bars represent the adjusted p-value of the GO term. **D**. (Top) Kaplan Meier survival curve with up to 5 year follow-up data for all samples grouped clinically assigned risk group (SR= standard risk, HR= high risk), ns= not significant by unpaired t-test. Black tick marks on the survival curve represents data that has been censored due to follow-up data <5 years. (Bottom) Kaplan Meier survival curve with up to 5 year follow-up data for all samples grouped by proteome cluster for clusters with >4 samples, **= p=value<0.01 by unpaired t-test. Black tick marks on the survival curve represents data that has been censored due to follow-up data <5 years.

We employed a data independent acquisition approach (DIA) using a spectral library of 10,130 proteins to quantify 8,590 proteins (Supplementary Fig. S5, Supplementary Tables S6 &S7). To determine if the proteome distinguishes leukemia types, non-cancer monocytes and cell lines, we first filtered for proteins with highly variable protein abundance across samples (Supplementary Table S8). Samples clustered distinctly by B-ALL, T-ALL and non-cancer monocytes and cell lines (Fig. 3B). In line with earlier reports, B-ALL cell lines cluster away from primary samples (12,13) suggesting phenotypical differences, thus highlighting the importance of direct studies of primary samples. Interestingly, there was one cluster comprised of several different sample types. Upon further investigation of blast percentage, we found this cluster to consist entirely of low-blast samples (the three non-cancer BM specimens, two low-blast T-ALL samples and one low-blast B-ALL sample). These findings demonstrate the sensitivity of our proteomics analysis to identify biological differences between sample types and perform unsupervised classification.

**Figure 3:**
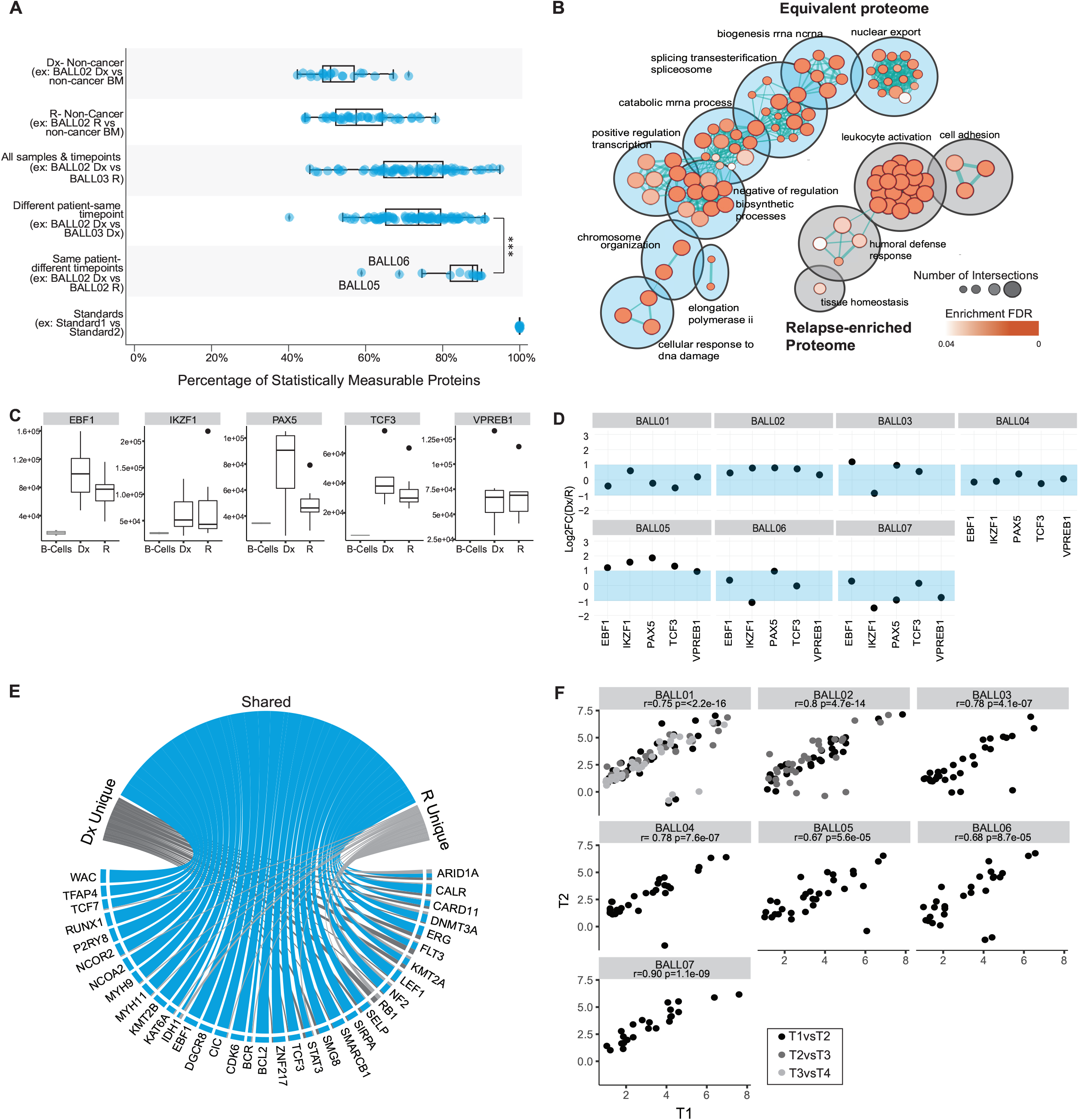
Cancer-associated proteins and processes remain stable through disease progression. **A**. Summary of tests for equivalence (Two-one-sided t-test (TOST) for equivalence, boundaries between log2FC<-1 and log2FC>1) of protein abundance between different groups and pairings. Only statistically measurable proteins are represented. Each dot represents the mean equivalence or difference of all protein abundance for a pairing. Significance is assigned by mann-whitney wilcoxon test, *** indicates p-value<=0.001. **B**. Pathway enrichment analysis of the stable population of proteins represented in blue and relapse enriched population of proteins represented in grey. The color of the circles indicates the enrichment FDR and size represents the number of identifications for the term. **C**. Abundance of transcription factors of interest for each sample separated by timepoint (T1 or T2, black) compared to protein expression in mature B-cells (n=2 samples) isolated from peripheral blood mononuclear cells (grey). **D**. Dot plots represent the log2FC of timepoint 1(T1)/timepoint 2(T2) for each of the proteins for each sample. The shaded blue area indicates the stable range of −1 to 1FC. *For patients with multiple timepoints (BALL01 and BALL02) only the log2ratio of the earliest timepoint/the latest timepoint is represented for simplicity. **E**. From the list of 269 pediatric cancer associated proteins (CAPs), 141 proteins were detected in our data and 45 proteins were deemed significant (LIMMA analysis of Initial Diagnosis (Dx) samples vs. non-cancer bone marrow (BM) samples and Relapse (R) vs non-cancer BM samples (log2FC>1, p-value adjusted BH-FDR<0.05). Circos plot summarizes significantly overexpressed cancer associated proteins (CAPs). **F**. Of the 45 proteins that were overexpressed, the protein expression for each protein that was overexpressed at Dx, was plotted as timepoint 1 (T1) vs timepoint 2 (T2), where T1 is the earliest timepoint available for the specimen (calculated as the log2(protein expression/the average protein expression in the non-cancer BM)). Pearson’s r correlation was calculated for all sample pairs. In cases that we had multiple time-points (BALL01 and BALL02) the correlation was calculated for consecutive pairings and are represented by the different colored dots.

To better characterize our largest patient group, we conducted a focused analysis of the samples in the two B-ALL clusters. Unsupervised hierarchical clustering of proteins with high variability in B-ALL (Supplementary Table S8) resulted in seven proteome clusters (Fig. 2C). Paired samples cluster closely together for four of six patients that had multiple timepoints (Fig. 2C) indicating high similarity consistent with our genomic findings. As well, some cytogenetic subtypes showed stronger trends in co-clustering, for example, cluster P3 primarily contained ETV6-RUNX1 patients and the hypodiploid patients clustered closely together although spread across two clusters. Cluster P4 was the largest cluster and consisted almost entirely of BCP-ALL or “other”, indicating that, although these samples are not characterized by a major shared genome alteration, the proteomes are remarkably similar. The remaining clusters were a mixture of subtypes, suggesting phenotypic similarities across cytogenetic subtypes (Fig. 2C). Gene ontology enrichment identified distinct biological processes with differential protein abundance between the clusters (Fig. 2C, Supplementary Table S9). Notably clusters P5, P6 and P7 have higher abundance of proteins involved in antigen presentation and leukocyte activation (cluster P3). Cluster P5 was enriched for processes related to actin and cytoskeleton organization while cluster P2 had the highest abundance in proteins relating to humoral immune response.

High-risk cases were associated with all clusters but enriched in cluster P4. Interestingly, stratification by risk group did not yield significant differences in 5-year event-free survival (event=relapse or death) (Fig. 2D). We next asked if stratification by unsupervised proteome-cluster showed any differences in 5-year event-free survival. Kaplan Meier analysis of the major proteome-clusters showed significant differences between clusters P2,3,7 and cluster P4 which was associated with a high event rate (Fig. 2E). Overall, our findings indicate phenotypic differences that are not solely linked to the common ALL cytogenetic subtypes, and highly similar proteomes between paired patients, consistent with our observation of genomic stability.

### Cancer-associated proteins and processes remain stable through disease progression

To interrogate the apparent similarity observed in patient-matched progression samples, we further examined the proteomes for matched Dx-R or R-R biopsies from 6 patients (BALL01, BALL03, BALL04, BALL05, BALL06, BALL07) plus one additional PDX-expanded Dx-R-R set (BALL02) (Supplementary Fig. S6 & S7, Supplementary Tables S10 & S11). To better understand inter- and intra-patient stability among the disease states, we tested all possible patient and timepoint pairings for statistically significant equivalence. As expected, the rate of significantly equivalent proteins was lowest when proteomes of non-cancer specimens were compared to proteomes of cancer specimens (63% or 68% equivalent to Dx or R, respectively) (Fig. 3A, Supplementary Fig. S8A). In contrast, >90% of robustly quantified proteins showed equivalent abundance when comparing proteomes of matched diagnosis and relapse specimens, or multiple relapses, from the same patient (Fig. 3A, Supplementary Fig. S8B, Supplementary Table S12). Consistent with our analysis of genomic stability and proteome cluster analysis (Fig. 2C), only BALL05 and BALL06 showed low equivalence (59% and 69% respectively) (Fig. 3A). At only 75% (median equivalence), diagnosis or relapse samples obtained from different patients show significantly lower proteome equivalence than matched samples from individual patients through progression (Fig. 3A).

To determine processes that are particularly stable throughout progression, we next performed a gene set and pathway enrichment analysis. Proteins found to be equivalent between cancer and non-cancer were removed prior to the enrichment analysis to eliminate ‘housekeeping’ mechanisms that are generally stable. Pathway enrichment analysis identified processes linked to overall cell survival as equivalent amongst cancer proteomes, including transcription related processes, metabolic processes, and cellular responses to DNA (Fig. 3B and Supplementary Table S13). Within the small set of proteins that differed between diagnosis and relapse, no enriched terms specific to the diagnostic timepoint could be identified. At relapse, processes involving cell adhesion, indicating more motility and proliferative mechanisms, and immune response (leukocyte activation, neutrophil activation involved in immune response, granulocyte activation, and exocytosis, among others) were found to be enriched (Fig. 3B and Supplementary Table S14). We next probed proteins involved in B-cell development, such as transcription factors IKZF1, EBF1, PAX5, VPREB1, and TCF (14) (Supplementary Table S15), that are commonly dysregulated in ALL. Overall, abundance of these proteins was significantly higher in cancer samples than in mature B-cells isolated from non-cancer peripheral blood mononuclear cells (PBMCs) (Fig. 3C). Moreover, for most patients, stable abundance was observed between disease states (Fig. 3C, D).

Finally, we identified 45 cancer-associated proteins (CAPs) to be significantly more abundant in diagnosis specimens (n = 6) or relapse specimens (n = 12) compared to non-cancer controls (n = 3) (Supplementary Fig. 9A-D and Supplementary Table S16), including several proteins that are commonly overexpressed in acute lymphoblastic leukemias such as FLT3, CDK6, and EBF1 (15). Given that the 45 CAPs are significantly more abundant in the ALL specimens than in the non-cancer specimens, they are likely linked to tumorigenic processes in our samples and would be of interest to determine their stability through disease progression. The majority (n = 36 proteins) showed increased abundance at both disease time-points (Fig. 3E) and their abundance levels were positively correlated between diagnosis and relapse (or relapse-relapse) (Pearson’s r = 0.75, p<2.2e-16); similarly, proteins with lower abundance (n = 10 proteins in either disease state) showed significant positive correlation between disease states (Pearson’s r = 0.70, p<3.5e-15) (Supplementary Table S17 and Supplementary Fig. S9E-F). Restricting the comparisons to matched specimens (n = 7 patients) confirmed that the high correlation was retained at the level of individual patients (Fig. 3F, Pearson’s r = 0.67 - 0.90), indicating the stability observed through disease progression across the global proteome is also observed when restricting the analysis to significantly more abundant CAPs.

### Whole proteome discovery-driven analysis identifies pan-ALL protein targets

The measured cytotoxicity was disappointing for agents informed by the Pediatric Match genetic variant-agent prioritization strategy (Fig. 2C) leading us to probe our proteome datasets for new targets. To discover pan-ALL protein targets, we first filtered for proteins identified in more than 40% of specimens and with a high overall abundance (log10 intensity) and strong abundance increase over non-cancer (log2 FC) (cut-off: at least 95^th^ percentile for both metrics) (Fig. 4A). We defined stable protein abundance between paired diagnostic and relapsed samples as a model variable for target discovery; using this criterium, and representing patient BALL01 as an example (Fig. 4B), we generated a ranked list of new pan-ALL targets, which included HSPB1, PARP1, and PRDX1 as top-ranked candidates (Fig. 4C). We selected to further characterize PARP1 as a candidate target since PARP1/2 inhibitors are already developmental therapeutics for a variety of pediatric tumors (16). Additionally, PARP1 is activated by DNA damage as a repair mechanism (17), and “cellular responses to DNA damage”, showed enrichment in our prior pathway enrichment analysis, providing further confirmation that this pathway is overexpressed and stable (Fig. 3E).

**Figure 4:**
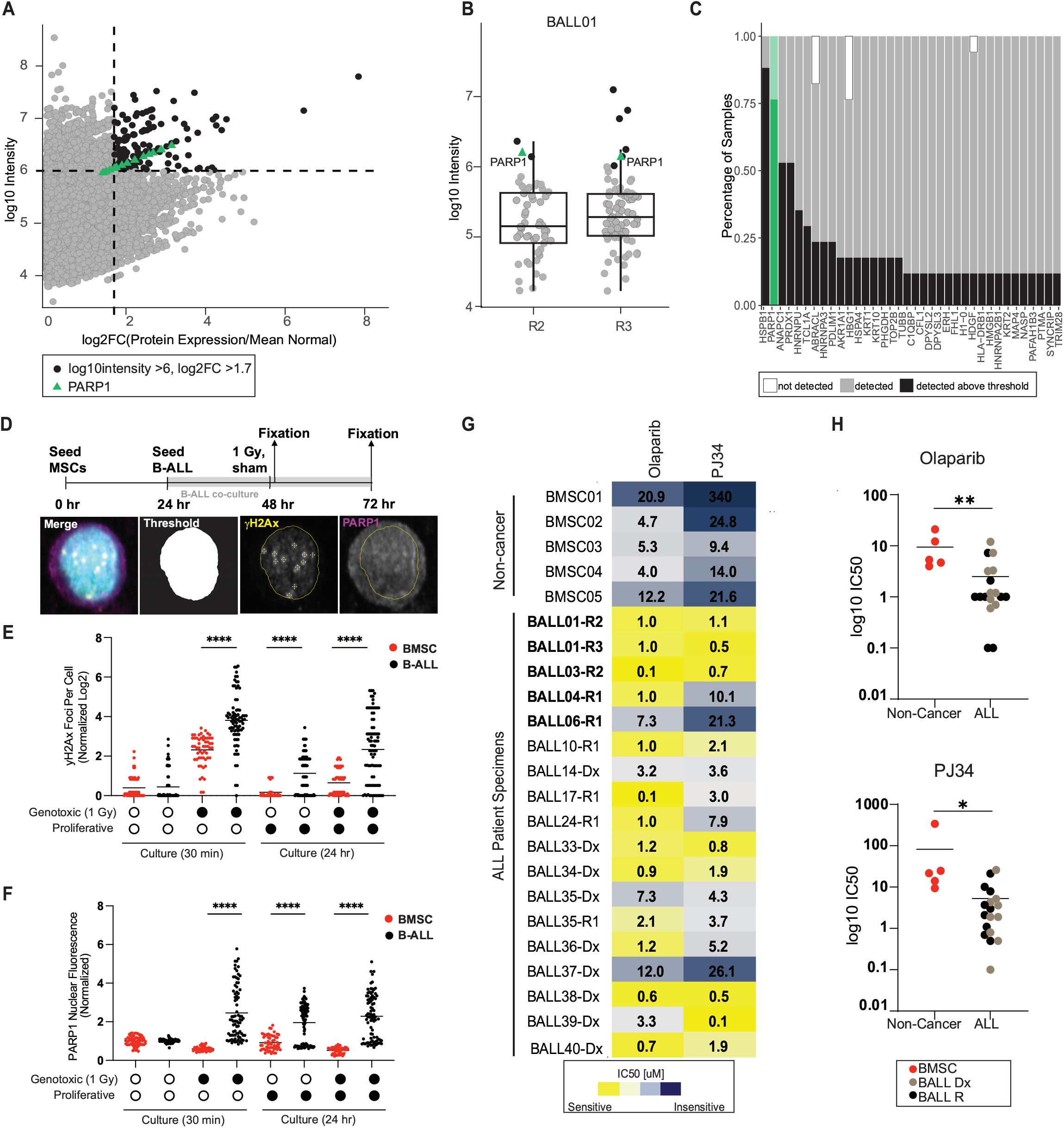
Whole proteome discovery-driven analysis identifies pan-ALL protein targets. **A**. Correlation of log2 fold-change (FC)/non-cancer vs protein expression (log10 intensity) for all proteins in samples from the paired Dx-R dataset (filtered for proteins identified in at least ten of seventeen samples). Dashed lines represent cut-offs for top five percent of the population for each parameter. Proteins that meet both cut-offs are black and PARP expression is represented by green triangles. **B**. Representative figure (BALL01) showing all proteins that are >log2FC of 1.7. Only the proteins that have log10 intensity >6.0 (top 5%) are in green. **C**. Summary of all proteins of interest plotted by percentage of samples the protein meets the indicated parameters (black), was identified but not meeting the parameters (grey) or not detected (white). PARP1 is highlighted in green. **D**. Experimental timeline and protocol (above) with image analysis pipeline (below) for primary B-ALL and BMSC cocultures followed by immunofluorescence analysis to quantify yH2Ax foci per cell and PARP1 nuclear fluorescence. **E**. Log2 yH2Ax foci per cell normalized to sham treatment at 30 minutes, quantified from immunofluorescence analysis of 2 BMSC (red) samples and 3 B-ALL (black) samples (n=30 cells per sample). Samples were treated with 1 Gy X-irradiation or sham conditions, and co-cultured with hTERT-MSCs for 30 minutes or 24 hours after treatment. (****p<0.0001, Welch’s t-test) **F**. Average PARP1 nuclear fluorescence per cell normalized to sham treatment at 30 minutes, quantified from immunofluorescence analysis of 2 BMSC (red) samples and 3 B-ALL (black) samples (n=30 cells per sample). Samples were treated with 1 Gy X-irradiation or sham conditions, and co-cultured with hTERT-MSCs for 30 minutes or 24 hours after treatment. (****p<0.0001, Welch’s t-test) **G**. Measured IC50 values for Olaparib or PJ34 measured against ALL or non-cancer samples from patients treated at BCCH (n=5 non-cancer, 8 diagnostic samples, 10 relapse samples). Samples were co-cultured with hTERT-MSCs and separately treated with the two PARP1/2 inhibitors. IC50 [µM] are colored by most sensitive in yellow to least sensitive in blue. Measurements represent the mean of n=2 replica wells from a single experiment. Bolded Patient IDs indicate patient samples analyzed in the pan-ALL target proteomic analysis. **H**. Dot plots for IC50 [µM] values for Olaparib or PJ34 measured against non-cancer specimens (red), primary diagnostic samples (light brown) or primary relapse samples (black). Measurements represent the mean of n=2 replica wells from a single experiment. Significance (*, **) is assigned by unpaired student’s t-test p-value <0.01 and p-value <0.001 respectively.

To validate the hypothesis that PARP1 elevation is reflective of an increased dependency on DNA repair in response to DNA damage, we examined the cellular response to genotoxic stress in viably-frozen ALL cells (n=3 patients) or non-cancer bone marrow-derived stem cells (BMSC) (n=2 donors). We co-cultured primary cells on hTERT-MSCs for 24 hours without (sham) or following exposure to genotoxic ionizing radiation (1 Gy X-ray) (Fig. 4D). After 30 minutes or 24 hours to allow induction or resolution of damage respectively, expression of the DNA damage marker gamma-H2AX or PARP1 was examined by immunofluorescence and the intensity of staining was normalized to the baseline levels measured at 30 minutes in sham treatment (Supplementary Fig.10A). This analysis revealed an expected increase of gamma-H2AX foci (DNA damage) 30 minutes after X-radiation in both BMSC and B-ALL samples (Fig. 4E, Supplementary Fig. 10B). The number of gamma-H2Ax foci was significantly elevated at 30 minutes after 1Gy X-radiation in B-ALL samples relative to control BMSCs (Fig. 4E) potentially indicating their hypersensitivity to genotoxic stress. The number of gamma-H2AX foci was reduced in both populations by 24 hours following X-radiation (Fig. 4E), albeit to a lesser extent in B-ALL cells, indicating either repair or clearance of damaged cells. However, the pattern of PARP1 expression was distinct for B-ALL cells relative to BMSC, increasing significantly in response to X-radiation (measured at 30 minutes) as well as following proliferative stress (at 24 hours in sham) (Fig. 4F)(Supplementary Fig. 10C), suggesting a reliance on PARP1 expression for B-ALL cell survival following stress.

To test a possible reliance on PARP1 for survival and its potential suitability as a target for therapeutic intervention we treated primary patient specimens with graded doses of two PARP1/2 inhibitors, Olaparib and PJ34. We sourced viably-frozen ALL samples (n=18) from the BCCH Biobank, including matched specimens used in our discovery cohort (n=4), additional ALL specimens (n=13), and non-cancer pediatric stem cell samples (n=5). Image-based drug screening of ALL cells co-cultured with MSC demonstrated high cytotoxic specificity of PARP1/2 inhibitors for ALL relative to non-cancer BMSC samples, as determined by IC50 values for each sample (Fig. 4G,H). The significant increase in sensitivity of ALL cells to PARP1/2 inhibitors relative to non-cancer cells indicates that this may be a potential pan-ALL therapeutic target that was initially discovered through protein abundance analysis.

## DISCUSSION

Molecularly guided targeted therapies have a high potential to improve outcomes for pediatric cancer patients. Yet, only 3-58% of patients receive molecularly guided therapies and even fewer report a positive response to treatment in NGS-guided trials (18). These unsatisfactory outcomes can be ascribed to multiple limitations, including the reliance on genomics for target identification, which cannot capture the plasticity of downstream transcriptional, translational and post-translational processes that impact target abundance and drug sensitivity, and the restricted enrollment for high-risk or relapsed cancers, which often progress quickly. Several initiatives, including ZERO and INFORM, have recently exemplified the use of precision oncology at diagnosis to help refine, or even change subtype diagnosis for several cancer types, which can lead to more appropriate treatment options (19,20). It is actively debated whether initiating molecular analyses for precision oncology at diagnosis is beneficial(18). Here, we advance the debate by contributing new evidence of high retention of potential drug targets in pediatric ALL. We show that the stability extends to and may even be more pronounced at the protein level and that proteome analysis can inform target selection in addition to and independent of genomic analysis.

One challenge to initiating precision medicine at diagnosis is the prospect that the dominant relapse clones contain distinct mutations and unique drug sensitivities (3,21–23). While it is estimated that 37% of primary tumours retain druggable events at relapse (18), we show that a far higher fraction (64% of 80 patients) of primary ALL retain at least one potential drug target. This high level of persistence was further reflected in the overall correlation of drug sensitivities between diagnosis and relapse. In our assay, genome variants predicted sensitivity to a targeted MEK inhibitor, which is consistent with prior studies (24,25). However, we found no correlation between the presence of *CDKN2A* deletion in primary ALL samples and sensitivity to CDK4/6 inhibition. Indeed, the utility of *CDKN2A* deletion to act as a predictive biomarker for sensitivity to CDK4/6 inhibitors is currently unresolved (26–29). Taken together, our data supports the notion that common genomic biomarkers are not sufficient to predict tumor sensitivity to variant-targeted monotherapies (30).

To supplement NGS-guided target identification, some precision oncology initiatives are complementing genome analysis with additional molecular strategies such as RNA-seq or Methyl-seq (18,19,31). Our study is among the first to conduct comprehensive targeted and non-targeted, paired Dx and Relapse proteome analyses of pediatric ALL. Proteome based clustering identified clusters that were independent of clinical features such as cytogroup and risk group. Instead, we identified a cluster of samples that is associated with poor 5-year EFS that primarily consisted of BCP-ALL or “other” sub-types. Paired Dx-R specimens also clustered closely together, and statistical testing for equivalence between paired samples confirmed this observation. This observation further adds to the emerging notion that proteome based molecular subtyping has the potential to identify larger groups that are more reflective of the actionable phenotype(32).

Combining tumor proteome insights with genomic data enables an even deeper understanding of disease progression. For example, targeted sequence analysis indicated considerable evolution in BALL03, but the CAP proteome analysis was highly stable (r=0.78 p<4.1e-07). Similarly, the relapse BALL04 sample gained a TP53 mutation, but CAP stability, proteome stability (equivalence of 90%), and drug responses were highly correlated with the diagnostic sample. Thus, the evolution of minor clones through disease progression may not dramatically impact the expressed proteomes.

Proteomic analysis can also reveal new potential targets (33–35). Consistently, we identified PARP1, PRDX1 and HSPB1 as overexpressed pan-ALL target candidates. The association of PARP1 with DNA damage and stress response was confirmed *in vitro* following exposure to genotoxic (X-radiation) and proliferative (24-hour co-culture) stress. Mechanisms to overcome such stressors enable leukemic cells to maintain active cell proliferation and prolong cell survival (36–38). Moreover, we validated the sensitivity of B-ALL samples to PARP1/2 inhibition *in vitro*. These proteome-based pan-ALL targets may warrant further investigation of their actionability in pediatric ALL.

Our retrospective design was useful for studying matched Dx-R samples, which are relatively rare and difficult to predict prospectively. We note that the conclusions drawn from our proteome analyses and image-based drug screening are limited by the number of matched patient samples examined, the variety of genetic subtypes, and the ethnic diversity of the patients treated in our single site cohort. In addition, image-based drug screening of ex vivo ALL-MSC co-cultures may not accurately reflect responses in patients although it has been shown to capture leukemia-intrinsic differences in cell proliferation and survival and, in the case of venetoclax, ex vivo responses correlate with strong *in vivo* antileukemic activity (39). With these potential caveats in mind, the findings from this study demonstrate clear potential utility for prospective proteogenomic variant identification for the targeted treatment of pediatric relapse ALL to be initiated at first diagnosis.

## Data Availability

All data produced in the present study are available upon reasonable request to the authors

## ACKNOWLEDGEMENTS

This work was supported by the Michael Cuccione Foundation (MCF) and the BC Children’s Hospital Foundation through the Better Responses through Avatars and Evidence (BRAVE) Initiative. Salary support was provided by the MCF (CJL, GSDR, CAM, PFL), the Canada Research Chairs Program (PFL), the Michael Smith Foundation for Health Research Scholar Program (PFL) and the University of British Columbia (AL, EKE).

Affiliations of authors: Department of Pediatrics, University of British Columbia, Vancouver, BC, Canada (AL, MG, NMA, CJL, GSDR, CAM); Michael Cuccione Childhood Cancer Research Program, BC Children’s Hospital, Vancouver, BC, Canada (AL, MG, NMA, CJL, GSDR, PFL, CAM); Department of Pathology, University of British Columbia, Vancouver, BC, Canada (EKE, PFL).

We thank Pascal Leclair for contributions made through the BRAVE Initiative and technical assistance. We gratefully acknowledge the participation of the patients and families that made this study possible and the BC Children’s Hospital nurses and physicians and Biobank staff for their tremendous efforts in collecting and maintaining specimens. We also acknowledge our consultations with the Modeling node and Proteomics node of the Terry Fox Precision Oncology for Young People Translational Research Program.

## MATERIALS AND METHODS

### Patient samples & non-cancer controls

Patient specimens were collected by Biobank staff at BC Children’s Hospital. Samples were taken with informed consent during routine clinical care. Sample collection and experiments were performed as approved by the University of British Columbia Children & Women’s Research Ethics, and conformed with standards defined in the WMA Department of Helsinki and the Department of Health and Human Services Belmont Report.

Mononuclear cells containing leukemic blasts were isolated by Ficoll-Paque PLUS density centrifugation, viably frozen and preserved. Aliquots of patient samples, and patient clinical information were de-identified prior to release for this study. Leukemia samples were immunophenotyped at the clinical hematopathology laboratory using established ALL subtype-specific 10-colour flow cytometry panels according to clinical standard operating procedures. Patient bone marrow morphology was assessed by hematopathologists and cytogenetics studies were performed by clinical cytogeneticists. Upon receipt of the specimens, patient mononuclear cells were thawed at 37°C for 1-2 minutes, washed 1x in warm RPMI-1640 medium containing 10% fetal bovine serum (FBS, Invitrogen, Waltham, Massachusetts, USA) and washed 2x with PBS and stored as 5×10^5 – 1×10^6 cells per cell pellet.

Bone marrow stem cells (BMSC) from five non-cancer individuals were initially collected following routine procedures for bone marrow stem cell transplantation, and remaining material was stored viably with the BCCH Biobank. In addition, we obtained bone marrow mononuclear cells from one healthy individual (non-cancer BM). Finally, for analysis of mature B-cells, PBMCs from five patients that did not have any hematological malignancies that were in a similar age range were combined.

### DNA/RNA extraction and sequencing

DNA and RNA extraction were performed using an Allprep (Qiagen) workflow. Library preparation and targeted sequencing was performed using the Oncomine Childhood Cancer Research Assay (OCCRA) on an Ion Chef and Ion Torrent S5 platforms (Thermo Fisher Scientific) following the manufacturer’s protocols. OCCRA comprises 2,031 unique DNA-based amplicons to detect SNVs, and CNVs, and 1,701 RNA-based amplicons to detect unique fusion or structural variants (10). The average read depth for the OCCRA panel was 5×10^6 – 7×10^6 per sample for DNA and 1×10^6 - 2×10^6 for RNA. SNVs were retrieved with Ion Reporter software (version 5.2). Copy number measurements were retrieved with Ion Reporter software (version 5.2) for genes with >5 probes, including those that were validated for copy number gains as described elsewhere (10).

### St. Jude’s Hospital (SJH) data curation

Data was downloaded from www.stjuderesearch.org/site/data/relapsed-all in December 2018. For our analysis, we included patients with Dx-R1 progression. To determine gene mutations as Dx unique, Shared, of Relapse unique, we included genes listed as “rise” as a shared mutation and genes listed as “fall” as Dx unique (Supplementary Table S3).

### Cytotoxicity analysis of variant-predicted drug response in paired Dx-R ALL samples

hTERT-immortalized mesenchymal stromal cells (MSCs) were provided by D. Campana (St. Jude’s Hospital). hTERT-MSCs were seeded at 5,000 cells per well in 200 μL of RPMI-1640 medium containing 10% fetal bovine serum (FBS, Invitrogen) and 1μM hydrocortisone (Sigma) in a 96-well plate (Corning, Corning, New York, USA), 24 hours prior to seeding with primary B-ALL or non-cancer bone marrow stem cells. Primary samples were thawed at 37°C for 1-2 minutes, washed 1x in warm RPMI-1640 medium containing 10% FBS and washed 2x with PBS, and stained with DAPI CFSE stain (Invitrogen) used to distinguish the ALL cells from the hTERT-MSC cells. The media was removed before adding 5×10^4 B-ALL cells in 100 μL of AIM-V medium (Thermo-Fisher).

Drug dilutions were prepared at 2x the final concentration (1nM, 10nM, 100nM, 1VM, 20μM, and 30μM) and 100μl of each drug dilution was added to 100μl of primary cells in each well. Cells were incubated with the drugs for 72 hours at 37°C in a 5% (v/v) CO2 incubator. Drugs used in the study: Palbociclib, Trametinib, Ruxolitinib, and Vismodegib (Selleck Chemicals LLC, Houston, TX, USA). For PARP1/2 inhibitors, the drugs were prepared for final concentrations of 1nm to 100µM in 10-fold increments PJ34 (SelleckChem), 0.1nM to 10µM in 10-fold increments for Olaparib (SelleckChem).

After 72 hours, GFP viability dye was added and incubated at room temperature for 1 hour. The plate was analyzed by a high content image analysis system (ImageXpress Micro XL). Images were taken using a 40X 0.75 NA dry objective with the MetaXpress 5.0.2.0 software (Molecular Devices Inc) on the ImageXpress Micro XL epifluorescence microscope (Molecular Devices Inc). DAPI and GFP (green fluorescent protein) emissions were acquired simultaneously with a 505DCXR beam splitter (Dual-View; Optical Insights, LLC) with the optical filters for DAPI excitation or GFP emission, respectively. For the analysis of the proportion of living cells, images were taken once per site using 50-ms exposures, 2×2 binned resolution, with 100% of full lamp intensity for each channel, and 25 optical sections spaced 500 μm apart. Post-acquisition processing of images was performed using MetaXpress offline.

Viability was calculated by taking the mean of (DAPI and GFP double-positive (DP) cells)/ (DAPI single-positive cells) for each drug-treated well. To account for relative viability of the primary cells in the assay, the drug-treated viability was normalized to the calculated viability for the vehicle-treated (DMSO) cells. For PARP1/2 inhibitors, normalized viability was assessed by the summation of DP cells across four sites in drug-treated wells / summation of DP cells across 4 sites in DMSO-treated wells. IC50 concentrations were calculated in GraphPad PRISM version 9 (GraphPad Software, San Diego, CA, USA) with the method “log inhibitor concentration vs normalized response”.

### Protein extraction and LC-MS/MS acquisition

Unless otherwise stated, reagents were purchased from Sigma Aldrich (St. Louis, Missouri, United States). Pellets of 5×10^5 – 1×10^6 cells were lysed in 50µl buffer containing 1% SDS (Fisher BioReagents, Pittsburgh, Pennsylvania, United States), 1X Pierce protease inhibitor (Thermo Fisher Scientific) in 50 mM HEPES (pH 8.0), followed by 5 minute incubation at 95 °C and 5 minutes on ice. The sample was incubated with benzonase (EMD Millipore/Novagen, Massachusetts, USA) at 37 °C for 30 minutes to shear chromatin. Following benzonase treatment, each sample was reduced with 10mM Dithiothreitol (DTT) dissolved in 50mM HEPES pH 8.0 (37°C, 30 minutes) and alkylated with 40 mM Chloroacetamide (CAA) dissolved in 50mM HEPES pH 8.0 (30 minutes in the dark) and quenched in 40 mM DTT for 5 minutes at room temperature.

Lysates were cleaned using single-pot solid-phase-enhanced (SP3) bead technique (40) using hydrophilic and hydrophobic Sera-Mag Speed Beads (GE Life Sciences, Issaquah, Washington, United States). Proteins were bound to paramagnetic beads with 80% ethanol (v/v), incubated for 18 minutes at room temperature, and washed twice with 90% ethanol using magnetic isolation. Beads were then resuspended in 30μl 200 mM HEPES, pH 8.0, and incubated with sequencing-grade trypsin (Promega Madison, Wisconsin, United States) at 1:50 protein ratio for sixteen hours at 37 °C, and afterwards acidified to pH 3-4 with formic acid. Peptide digests were de-salted on Nest Group Inc. C18 spin columns with 0.1% trifluoroacetic acid (TFA), eluted with 60% acetonitrile in 0.1% FA and dried in a vacuum concentrator. Dried samples were resuspended in 0.1% formic acid (FA).

### Library Preparation-High pH Reverse-Phased Fractionation (for BCCH cohort 1)

Depending on final protein amount, 1-4ug of protein was taken from each sample and combined into one pool for fractionation. Fractionation was performed on a Kinetic EVO C18 column (2.1 mm x 150 mm, 1.7 µm core shell, 100Å pore size, Phenomenex) connected to an Agilent 1100 HPLC system equipped with a diode array detector (254, 260, and 280 nm). A flow rate of 0.2 ml per minute was maintained on a 60 min gradient using mobile phase A (10mM ammonium bicarbonate, pH 8, Fisher Scientific, cat. no. BP2413-500). Elution was with mobile phase B (acetonitrile, Sigma-Aldrich, cat. no. 34998-4L) from 3% to 35%. Peptide fractions were collected each minute across the elution window. A total of 48 fractions were combined to a final set of 24 (e.g fraction 1 + 25 as final fraction 1), and dried in a SpeedVac centrifuge. Peptides were resuspended in 0.1% FA in water (SC235291, Thermo Scientific) prior to mass spectrometry analysis.

Peptides were analyzed using a Thermo Scientific Easy-spray PepMap™RSLC C18 column (75 μm x 50 cm, 2 μm, 100Å; ES803), maintained at 50 °C on an Easy-nLC 1200 connected to a Q Exactive HF mass spectrometer (Thermo Scientific). Peptides were separated over a 3 h gradient consisting of Buffer A (0.1% FA in 2% acetonitrile) and 3 to 30% Buffer B (0.1% FA in 95% acetonitrile) at 250 nL/min. MS acquisition was performed with full scan settings between 400 and 1800 m/z, resolution of 60000, AGC target of 5 e4, and Maximum IT of 75 ms. Stepped collision energy (NCE) was 28. MS2 scan settings were as follows: isolation window of 1.4 m/z, AGC target of 5 e4, maximum IT of 50 ms, at resolution of 15,000 and dynamic exclusion of 20.0 s.

### Library Preparation-Gas Phase Fractionation (for BCCH cohort 2&3)

Online gas-phase fractionation was performed. 1-2 μg de-salted peptides from select samples were combined into a single pool and analyzed in ten fractions, 1 μg per fraction. The first eight fractions (340 m/z to 760 m/z) were analyzed over a 60 m/z window (ie 340 m/z – 400 m/z is fraction 1) each with a loop count of 30 and window size of 2 m/z. The final two fractions (820 m/z-1180 m/z) were analyzed over 180 m/z window each, with a loop count of 30 and 6 m/z window.

Peptides from cohort 1 were separated on a Thermo Scientific Easy-spray PepMap™RSLC C18 column (75 μm x 50 cm, 2 μm, 100Å; ES803) and from cohort 2&3 were separated on a PharmaFluidics 50cm uPAC™ (ESI Source Solutions, Woburn, MA, United States), maintained at 50 °C on an Easy-nLC 1200 connected to a Q Exactive HF mass spectrometer (Thermo Fisher Scientific). The peptides were separated over a 3 hour gradient consisting of Buffer A (0.1% FA in 2% acetonitrile) and 2% to 80% Buffer B (0.1% FA in 95% acetonitrile) at 300 nL/min. MS acquisition consisted of a MS1 scan ranges specified above for each fraction (AGC target of 3e6 or 60 ms injection time), and resolution of 120,000. DIA segment spectra were acquired with a AGC target 3e6, resolution 30,000, auto ms injection time, and stepped collision energy of 25.5, 27, 30. The library was also supplemented with DIA data from each individual sample.

### Sample MS acquisition

1 μg of peptides was injected for analysis for each sample. Samples were randomized and we obtained duplicate injections (cohorts 2&3) when possible. The DIA method consisted of a MS1 scan from 300 to 1650 m/z (AGC target of 3e6 or 60 ms injection time), and resolution of 120,000. DIA segment spectra were acquired with a twenty-four-variable window format, (AGC target 3e6, resolution 30,000, auto for injection time), and stepped collision energy of 25.5, 27, 30. We added indexed retention time (iRT) peptides (Biognosys, Schlieren, Switzerland) to each sample for retention time normalization and quality control.

### Full BCCH Cohort-Proteomics Data Analysis

The BCCH cohort consisted of three sample sets. The spectral library for each of the three sample sets was combined in Spectronaut into one library of 10,130 proteins. All DIA files were searched together with the combined spectral library. Briefly, the raw DIA files were analyzed with Spectronaut Pulsar X (Biognosys, Schlieren, Switzerland) using a human FASTA file from UniProt (reviewed 20200309). This FASTA file includes common contaminants. Additionally, a FASTA file for iRT peptides, provided by Biognosys, was included in the search. Search was performed using the factory settings including specificity for Trypsin, Carbamidomethyl (C) as a fixed modification, and Acetyl (protein N-term) and Oxidation (M) as variable modifications. Precursor, and protein identifications false discovery rate (FDR) threshold was set to 1%, while the threshold for peptide was 0.5%. The data was normalized in Spectronaut based only on proteins identified in all samples and then further processed for batch effect removal by HarmonizR (41).We utilized HarmonizR for batch effect correction is that it allows the retention of the proteins that otherwise would be dropped due to containing missing values (Supplementary Fig. S5).

For the analysis of the full cohort, data was filtered for proteins identified in at least 25% of each cohort, and the remaining missing values were imputed using a “down-shifted normal” imputation strategy resulting in a total of 7307 proteins used for analysis. For hierarchical clustering we filtered for highly variable proteins by calculating the relative FC (protein intensity/median protein intensity) for each protein, and selected only the proteins with a log2FC>2 in at least 6 samples (3907 proteins).

The B-ALL analysis was restricted to proteins identified in all B-ALL samples (3857 proteins). Highly variable proteins with a log2FC>1 in at least 5 samples were selected (935 proteins). For gene ontology analysis we used g:Profiler (BIIT! Research Group, g:Profiler version e104_eg51_p15_3922dba, database updated on 07/05/2021) with an FDR threshold of 5%. To attain better resolution of the pathway visualization, we followed a similar strategy previously described (42); we limited the terms to GO: Biological Process (BP) and limited the number of intersections to 1000. The 5 terms with most significant adjusted p-value were selected for visualization.

### Survival Analysis

Kaplan Meier survival analysis was performed in GraphPad Prism. Patients that had less than 5 year follow up data were censored, meaning the sample was still included but the end of the follow up data is indicated on the survival curve. A long rank test for trend was used to determine significance.

### Paired Dx-R Cohort-Proteomics Data Analysis

For a summary of data quality and filtering, refer to Supplementary Fig. S4, S5. The data was analyzed in spectronaut as previously described. A minimum of 2 peptides were required for quantitation. Protein intensities were normalized in Spectronaut using the “global” setting, which normalizes by median protein intensity per sample. Duplicate injections were averaged (mean). A pool of samples was created as a “standard” that was injected periodically throughout the course of the sample run, to measure data reproducibility and monitor MS performance. Proteins that were quantified with >50% CV in the 10 standards were removed from all samples as they are presumed to be unreliably quantified. Furthermore, proteins identified in <10% of the samples were removed. Finally, missing values were imputed using a “down-shifted normal” imputation strategy. Briefly, a normal distribution was created out of the overall sample distribution, then shifted to lower values using magnitude of 3.5.

To summarize, we identified a total of 8153 unique proteins with an average of 6600 proteins per sample using a data independent acquisition approach (DIA) with a spectral library of 8183 proteins derived from gas-phase fractionated sample pool (Supplementary Fig. S6A). After quality assessment and data filtering (Supplementary Fig.6B), we quantified an average of 5100 proteins per sample with at least 2 peptides at less than 0.05% FDR.

### Fluorescence-activated cell sorting to isolate mature B-cells

To investigate comparison to B-cells, we included data from naïve and memory B-cells isolated by flow cytometry from age-matched PBMCs. To decrease processing and patient-specific variability and increase sorting efficiency, we combined PBMCs from five patients that did not have any hematological malignancies and were in a similar age range (see Supplementary Table S1). Briefly, we thawed 1 vial (approximately 5×10^6 – 10×10^6 cells per vial) and washed once with sterile FACS buffer (PBS (Thermo Fisher Scientific) + 2% FBS (Thermo Fisher Scientific)). We used 25×10^6 live cells for staining. Cells were pelleted and resuspended in approximately 100 µl of FACs buffer. We utilized a previously established 8-color flow panel designed to identify CD45+ lymphocytes and then optimally separate T-cells and B-cells and their respective subpopulations; for this experiment we aimed to isolate two different mature B-cell populations, naïve B-cells (CD45+CD3-CD19+CD10-CD20+CD27-). and memory B-cells (CD45+CD3-CD19+CD10-CD20+CD27+). The panel consists of CD45-AF488(HI30), CD3-BV510(UCHT1), CD19-APCFire(HIB19), CD4-AlexaFluor700(SK3), CD8-PECy7(RPA-T8), CD10-BV421(HI10a), CD20-APC(2H7), CD27-PE(O323) (BioLegend). We added BV staining cocktail for optimal performance of the BV dye and human FC blocker to reduce unspecific binding. Staining was performed for 20 minutes at 4 ^⸰^C in the dark. After staining, the cells were washed with FACS buffer, centrifuged at 1500 rpm and resuspended in 1 mL of FACS buffer. Just prior to sorting, we added 7AAD for viability. Sorting was done at the Center for Molecular Medicine and Therapeutics (CMMT) flow sorting core at BCCHR on an Astrios FACS sorter.

### Statistical Analysis

To look at protein stability, we established a hybrid method that compares robustly quantified proteins using equivalence and differential expression testing. We normalized the quantitative protein data using median centring and standard samples 1,2,3, and 9 were removed. To create a robust comparison between statistically different and equivalent proteins for a given pair of samples, we employed coefficient of variation (CV) based filtration to filter out proteins with unstable quantification. Proteins with >20% CV between the technical replica for each sample were removed. Only proteins that passed this threshold in each-pairwise comparison remained, therefore there were no missing values or imputed data. The statistical tests were done on each pair of sample comparisons with their two technical replicates. The differential expression testing was done using a ttest_ind function from the stats package. Proteins resulting with p-value < 0.05 and logFC > 1 were designated as statistically different. The equivalence testing was done using the TOST two_raw function from the TOSTER package setting logFC<-1 and logFC>1 as boundaries for equivalence.

To obtain a list of equivalent proteins for gene ontology enrichment, we first ensured the protein was equivalent in at least two of the seven Dx-R pairings. We next removed the proteins attributed to “housekeeping” functions by creating a cancer vs. non-cancer pairings equivalent list, which also required the protein was detected in at least two pairings. Housekeeping proteins were then removed from our paired Dx-R equivalence list. To obtain the “difference” list, we similarly required “difference” in protein expression in at least two of the seven Dx-R (or R-R) pairings. To determine if the protein was Dx or R enriched, we first summed the FC across the pairings and if the total FC was below −1 it was considered R enriched, above +1 was considered Dx enriched and in-between was counted for both Dx and R, leading to two lists for analysis (Dx+ both and R+both).

Gene ontology enrichment analyses was performed as previously described. For visualization of the enriched terms, we employed Cytoscape (43) (version 3.8.2) and utilized the “EnrichmentMap” and “AutoAnnotate” packages. For TF enrichment, we created a list of equivalent proteins as previously described. However, these proteins had to be identified in at least three of the paired samples, and at least three of the cancer vs. non-cancer pairings. The enrichment was obtained from gProfiler, as described above. The background list for all GO analyses was a list of all proteins quantified in the data set.

### Identification of cancer-associated proteins

Proteins were selected from two large-scale pediatric cancer studies that identified commonly mutated genes, genes of prognostic importance, and potential cancer drivers (9,10) (Supplementary Table S6). Additionally, targets were included from the OCCRA panel, which is similar to the Oncomine panels used in NCI-COG Pediatric MATCH precision medicine trial (21) (Supplementary Table S6).

For statistical analysis, the “Limma” package for R was used. P-value was adjusted using the Benjamin-Hochberg and the FDR threshold was 5%. Circos plots were created in R using the package “Circularize”; data was first filtered for proteins that were overexpressed relative to the non-cancer controls. Pearson correlation coefficient was calculated for each pairing based on proteins that were overexpressed in Dx.

### Primary Cell Irradiation

hTERT-MSCs were seeded at 70% confluency per well in 4mL of RPMI-1640 medium containing 10% fetal bovine serum (FBS, Invitrogen) and 1μM hydrocortisone (Sigma) 24 hours prior to seeding with primary B-ALL or stem cells from bone marrow. To seed primary cells, RPMI-1640 complete medium was removed before adding 3.2×10^6^ primary cells, recovered from cryopreserved samples, in 4mL of AIM-V medium. Both primary cells and hTERT-MSCs were incubated at 37°C in a 5% (v/v) CO2 incubator. After 24 hours co-culture, primary cells were removed from co-culture and treated with 1Gy X-irradiation or sham conditions. Primary cells were then added back to hTERT-MSC co-culture. Half of the primary cells were fixed 0.5hrs after irradiation, with the other half fixed 24 hours after irradiation. Cells were concentrated onto a slide using the Epridia Cytospin 4 centrifuge (Fisher) and fixed in methanol at −20°C for 5 minutes before storage at −20°C.

### Primary Cell Immunofluorescence

Cells were concentrated onto slides using the Epridia Cytospin 4 centrifuge (Fisher) and fixed in methanol at −20°C for 5 minutes. Cells were outline with a PAP pen (abcam) and blocked in PBS with 0.2% Triton X-100 and 3% BSA for 1 hour at room temperature. Antibodies were diluted in PBS with 0.2% Triton X-100 and 3% BSA. Primary antibodies were diluted (yH2Ax 1:500, PARP1 1:250) and incubated with slides overnight at 4°C. Cells were then washed three times in PBS. The slides were incubated with diluted secondary antibodies (Alexa Fluor 1:2000) at room temperature for 1 hour in the dark. Slides were washed three times in PBS and incubated with Hoechst stain for 15 minutes. Slides were then washed two times in PBS and coverslips were mounted with ProLong Gold Antifade (Invitrogen) reagent.

### Confocal Microscopy and Image Analysis

Fixed cells were imaged using the Fluoview software (Olympus) connected to the Olympus Fluoview FV10i confocal microscope. Image stacks of 5 optical sections with a spacing of 0.5 μm through the cell volume were taken using a 60X 1.2 NA oil objective. PARP1 stained with AlexaFluor 594 was imaged at 50% sensitivity, and 40% laser power. yH2Ax stained with Alexa Fluor 647 was imaged at 50% sensitivity, and 40% laser power. Hoechst nuclear stain was imaged at 40% sensitivity and 13% laser power. ImageJ v1.46j (National Institute of Health) was used to generate maximum intensity Z-projection of the fluorescent channels, and subsequent analysis. Nuclear masks were generated for each cell (Make Binary) and the resulting ROI (Analyze Particles) was used to identify the nuclear region of analysis for yH2Ax and PARP1 channels. yH2Ax foci per cell was quantified using the Find Maxima process (prominence > 750). PARP1 nuclear fluorescence was quantified using the Measure analysis. Statistical analysis was performed using GraphPad Prism with Welch’s t-test, as indicated in each figure. The results were considered statistically significant at P<0.05.

## Data Availability

MS data have been deposited to the Proteome Consortium (http://www.proteomexchange.org) via the MassiVE (https://massive.ucsd.edu/) partner repository data set MSV000091012.

## SUPPLEMENTARY FIGURE LEGENDS

**Supplementary Figure S1: Genomic stability in paired ALL specimens from the BCCH cohort**

Mutated gene products identified through targeted DNA/RNA-fusion sequencing of paired diagnosis (Dx) and relapse (R) samples. Mutations detected in both Dx and R samples are represented by blue boxes, mutations unique to Dx are dark grey, and R unique are light grey. CNVs are full boxes and SNVs are outlined boxes. The pie diagrams at the top summarize the number of mutations for each category for each patient. An asterisk indicates multi timepoint patients; BALL01 R2-R3-R4-R5-R5P, BALL06 Dx-R1-R2.

**Supplementary Figure S2: Summary of detected genomic lesions in paired ALL specimens from the BCCH cohort**

**A-K**. Line graphs for each patient treated at BCCH represent genomic lesions identified through targeted NGS in samples collected through disease progression. Only abnormal variants are plotted. CNVs are plotted by the number of copies detected and SNVs are plotted by the allelic frequency. Venn diagrams for each patient display variants that are unique to diagnosis (Dx unique), unique to relapse (R unique), or shared between samples in a progression series. An asterisk indicates the lesion is a gene-fusion detected by RNA analysis.

**Supplementary Figure S3: Summary of detected genomic lesions in paired ALL specimens from the SJH cohort**

**A**. Bar graphs for each patient with B-ALL treated at SJH (n=49) illustrate the fraction of variants that are unique to diagnosis (Dx Unique, dark grey bars), unique to relapse (R unique, light grey bars), or shared (blue bars), identified through whole genome sequencing in samples collected through disease progression. Venn diagrams are shown for selected patients to display the distribution of variants in a progression series.

**B**. Bar graphs for each patient with T-ALL treated at SJH (n=20) illustrate the fraction of variants that are unique to diagnosis (Dx Unique, dark grey bars), unique to relapse (R unique, light grey bars), or shared (blue bars), identified through whole genome sequencing in samples collected through disease progression. Venn diagrams are shown for selected patients to display the distribution of variants in a progression series.

**Supplementary Figure S4: Dynamics of genomic lesions detected in the SJH and BCCH cohorts**

**A**. Bar plot for the most frequent variants (top 17) detected in both Dx and paired R samples (shared) plotted as the fraction of samples containing the shared variant in each cohort (n=11 from BCCH (light pink), n=69 from SJH (red)). Variants were detected in the BCCH cohort through targeted NGS while variants were detected in the SJH cohort through whole genome sequencing. Thus, variants that cannot be detected in the BCCH samples with the targeting sequencing assay are indicated with ^.

**B**. Bar plot for the most prevalent genes (top 16) detected in the combined BCCH and SJH cohorts. Variants are categorized as shared (blue), Dx unique (dark grey) or R unique (light grey) and plotted as the fraction of occurrences. The total number of times the gene was identified is displayed on the right end of the bar.

**C**. Bar plot classifying variants (SNV, CNV, Fusion) as shared (blue), Dx unique (dark grey) or R unique (light grey) and plotted as the fraction of occurrence in the BCCH cohort.

**D**. Dot plot for fraction of shared variants versus time to relapse for 59 B-ALL patients (n=10 from BCCH represented by light blue circle, n=49 from SJH represented by dark blue circles).

**E**. Dot plot for fraction of shared variants versus time to relapse for 21 T-ALL patients (n=1 from BCCH represented by light pink circle, n=20 from SJH represented by red circles).

**Supplementary Figure S5: BCCH Proteome Full Cohort-Batch Correction and Data Quality**

**A**. (Left) PCA of proteins identified in all samples (2995) prior to batch correction. The points are colored by cohort; cohort 1 in purple, cohort 2 in yellow, and cohort 3 in green. (Right) Hierarchical clustering of the 2995 proteins prior to batch correction scaled by min/max. The dendrogram indicates the clustering of the samples and the color bar indicates the cohort the sample was from.

**B**. (Left) PCA of proteins identified in all samples (2995) after batch correction. The points are colored by cohort; cohort 1 in purple, cohort 2 in yellow, and cohort 3 in green. (Right) Hierarchical clustering of the 2995 proteins after batch correction scaled by min/max. The dendrogram indicates the clustering of the samples and the color bar indicates the cohort the sample was from.

**C**. Visual of data completeness in all of the samples after batch correction. Percentage of completeness is represented in blue and missingness is represented in grey.

**Supplementary Figure S6: Description of proteomic data filtering pipeline and quality assessment**

**A**. Total protein groups identified in each sample, prior to any filtering. The category of the sample type is listed across the top of each group.

**B**. Diagram of the filtering workflow to attain the final proteomics dataset utilized for subsequent analyses.

**C**. Upset plot for ten of the eleven standards (standard nine was removed due to a clear technical issue). Numbers reported are based on protein groups quantified by a minimum of two peptides at a precursor q-value threshold of 0.5% FDR.

**D**. The cv for protein quantity across the ten standards was assessed and plotted by percent coefficient of variation (CV). The darkest purple bar at the bottom represents the number of proteins with a CV of less than 10% and so on, with the lightest bar at the top representing the number of proteins with a cv greater than 50%. This fraction of proteins with cv >50% was removed from the remaining data with the assumption these are unstably quantified between samples.

**E**. A violon plot demonstrating the median CV of protein quantification across the ten standards (including those >50% CV). Dashed line indicates the median CV (15.7%).

**Supplementary Figure S7: Evaluation of individual sample data quality**

**A**. Bar plot to demonstrate data completeness between replica of each sample. Proteins that were identified in both replica are represented in black, dark grey represents proteins that were only identified in one of the replica, and light grey represents proteins that were entirely missing from the pair.

**B**. CV between replica is represented as described in panel S5 D; The darkest color bar at the bottom represents the number of proteins with a CV of less than 10% and so on, with the lightest bar at the top representing the number of proteins with a cv greater than 50%.

**Supplementary Figure S8: Statistical analysis of paired samples**

**A**. Summary of tests for differential expression and equivalence between different groups and pairings. Proteins that are statistically equivalent are represented in blue (Two-one-sided t-test (TOST) for equivalence, boundaries between log2FC<-1 and log2FC>1), proteins that are statistically different are represented in grey (student’s ttest p-value<0.05, log2FC>1). The bar represents the mean equivalence or difference of all protein expression for each pairing within the group.

**B**. Similar representation for each individual patient pairing of the group “Same patient-diff timepoints” group. The numbers on the right of each bar indicate how many statistically measurable proteins were in each pairing.

**Supplementary Figure S9: Significant cancer associated proteins (CAPs)**

**A**. 141 proteins were tested using LIMMA (log2FC>1, p-value adjusted FDR<0.05); Initial Diagnosis (Dx) samples vs. Non-cancer BM samples on the left and Relapse (R) vs Non-cancer BM samples on the right. Proteins that are significantly over expressed in both Dx and R are colored in blue and those that are unique to either Dx or R are colored in grey.

**B**. From the list of 269 pediatric cancer driving proteins (black circle), 141 proteins were detected in our data (gray circle) and 45 proteins were deemed significant (red circle).

**C**. Sixteen of the overexpressed proteins were shared between Dx and R (bottom)

**D**. Of the 45 proteins that were overexpressed at Dx, the protein expression for each protein at each timepoint was plotted as timepoint 1 (T1) vs timepoint 2 (T2) (calculated as the log2(protein expression/the average protein expression in the non-cancer bone marrow (BM) samples). A Pearson’s r correlation was calculated for the entire dataset.

**E**. Of the 45 proteins that were under-expressed at Dx, the protein expression for each protein at each timepoint was plotted as timepoint 1 (T1) vs timepoint 2 (T2) (calculated as the log2(protein expression/the average protein expression in the non-cancer bone marrow (BM samples). A Pearson’s r correlation was calculated for the entire dataset.

**Supplementary Figure S10 Summary of PARP1 and yH2Ax immunofluorescence data**

**A**. Representative image showing immunoflourescence staining of yH2Ax and PARP1 individually, and merged with Hoechst nuclear stain for primary sample BALL03-R2. Samples were treated with 1 Gy X-irradiation or sham conditions, and co-cultured with hTERT-MSCs for 30 minutes or 24 hours after treatment.

**B**. Log2 yH2Ax foci per cell normalized to sham treatment at 30 minutes, quantified from immunfluorescence analysis of 2 BMSC (red) samples and 3 B-ALL (black). Individual primary samples are indicated by number (1=BMSC02, 2=BMSC05 3= BALL04-R1, 4=BALL01-R2, 5=BALL03-R2) (n=30 cells per sample). Samples were treated with 1 Gy X-irradiation or sham conditions, and co-cultured with hTERT-MSCs for 30 minutes or 24 hours after treatment.

**C**. Average PARP1 nuclear fluorescence per cell normalized to sham treatment at 30 minutes, quantified from immunfluorescence analysis of 2 BMSC (red) samples and 3 B-ALL (black). Individual primary samples are indicated by number (1=BMSC02, 2=BMSC05 3= BALL04-R1, 4=BALL01-R2, 5=BALL03-R2) (n=30 cells per sample). Samples were treated with 1 Gy X-irradiation or sham conditions, and co-cultured with hTERT-MSCs for 30 minutes or 24 hours after treatment.

