## Supplemental Figures S1-S10 for "Persistence of targetable lesions, predicted therapy sensitivity and proteomes through disease evolution in pediatric acute lymphoblastic leukemia"

**Supplementary Figure S1: Genomic Stability in paired ALL specimens from the BCCH Cohort**

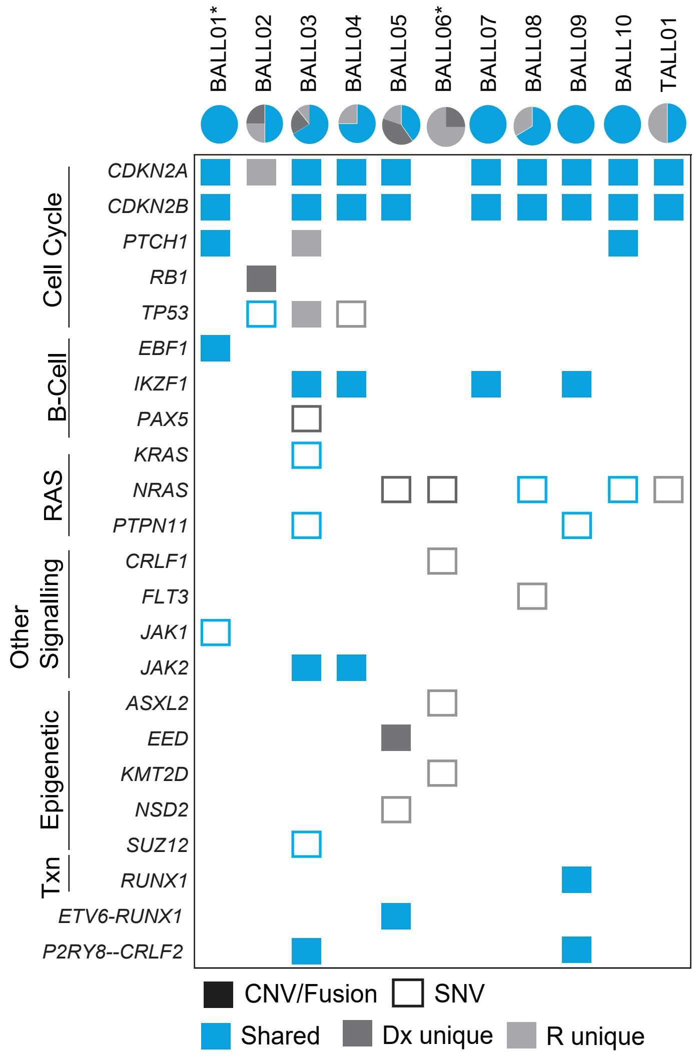


**Supplementary Figure S1: Genomic Stability in paired ALL specimens from the BCCH Cohort**
Mutated gene products identified through targeted DNA/RNA-fusion sequencing of paired diagnosis (Dx) and relapse (R) samples. Mutations detected in both Dx and R samples are represented by blue boxes, mutations unique to Dx are dark grey, and R unique are light grey. CNVs are full boxes and SNVs are outlined boxes. The pie diagrams at the top summarize the number of mutations for each category for each patient. An asterisk indicates multi timepoint patients; BALL01 R2-R3-R4-R5-R5P, BALL02 Dx-R1-R2.

Supplementary Figure S2: Genomic lesions detected in paired ALL specimens from the BCCH cohort

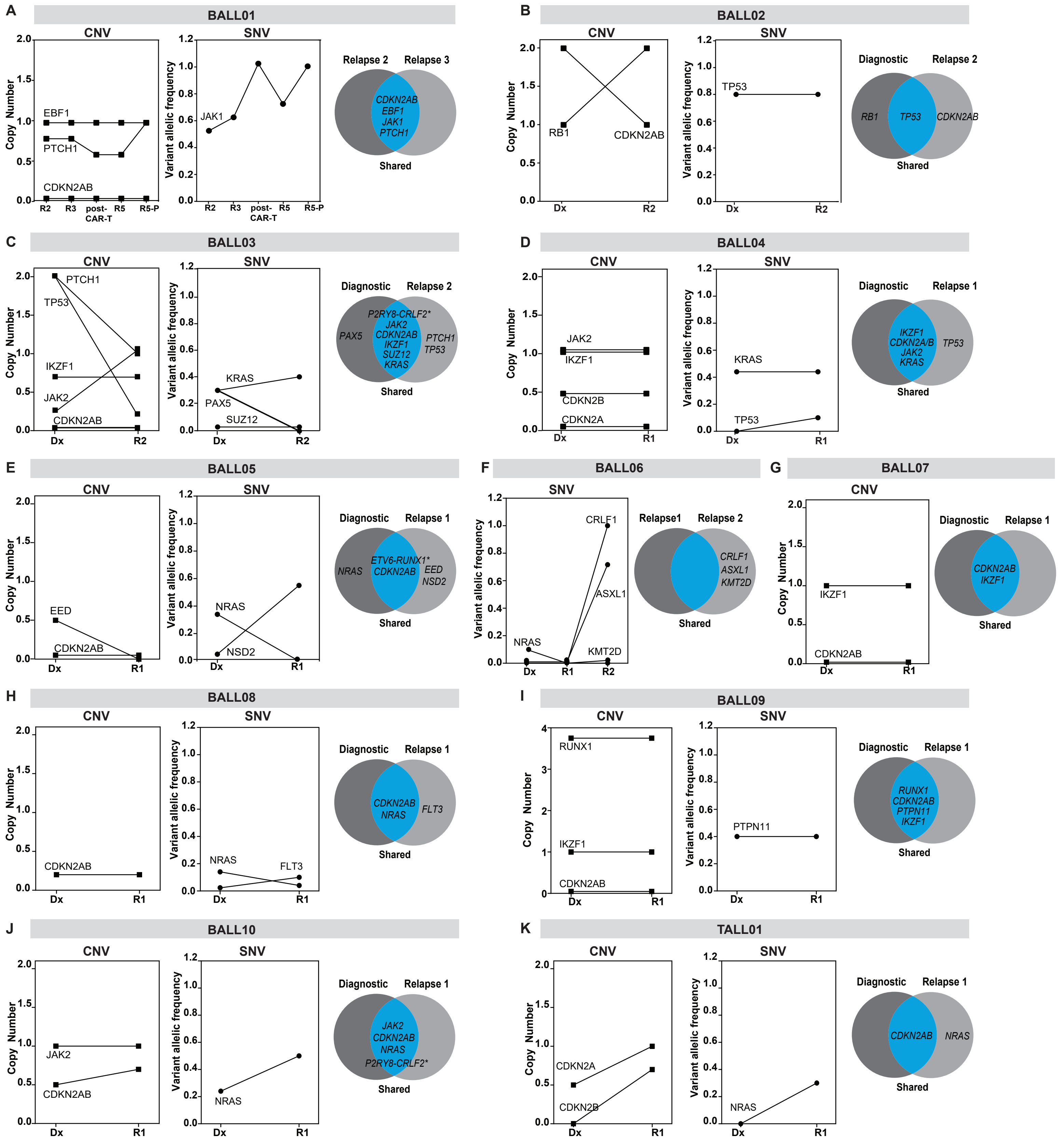


**Supplementary Figure S2: Genomic lesions detected in paired ALL specimens from the BCCH cohort**
**A-K.** Line graphs for each patient treated at BCCH represent genomic lesions identified through targeted NGS in samples collected through disease progression. Only abnormal variants are plotted. CNVs are plotted by the number of copies detected and SNVs are plotted by the allelic frequency. Venn diagrams for each patient display variants that are unique to diagnosis (Dx unique), unique to relapse (R unique), or shared between samples in a progression series.

Supplementary Figure S3: Genomic lesions detected in paired ALL specimens from the SJH cohort

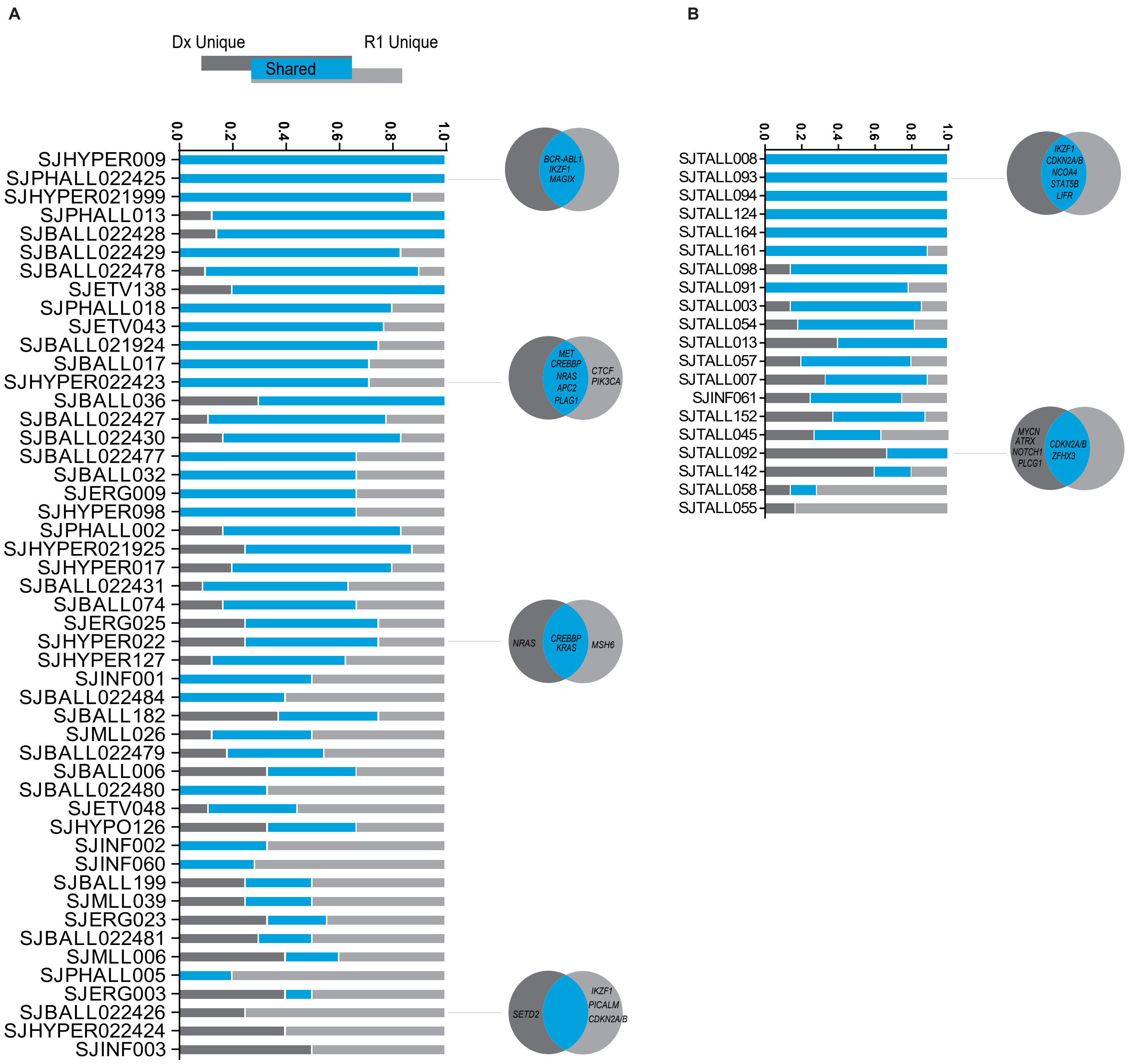


**Supplementary Figure S3: Genomic lesions detected in paired ALL specimens from the SJH cohort**

A. Bar graphs for each patient with B-ALL treated at SJH (n=49) illustrate the fraction of variants that are unique to diagnosis (Dx Unique, dark grey bars), unique to relapse (R unique, light grey bars), or shared (blue bars), identified through whole genome sequencing in samples collected through disease progression. Venn diagrams are shown for selected patients to display the distribution of variants in a progression series.

B. Bar graphs for each patient with T-ALL treated at SJH (n=20) illustrate the fraction of variants that are unique to diagnosis (Dx Unique, dark grey bars), unique to relapse (R unique, light grey bars), or shared (blue bars), identified through whole genome sequencing in samples collected through disease progression. Venn diagrams are shown for selected patients to display the distribution of variants in a progression series.

Supplementary Figure S4: Dynamics of genomic lesions detected in the SJH and BCCH cohorts

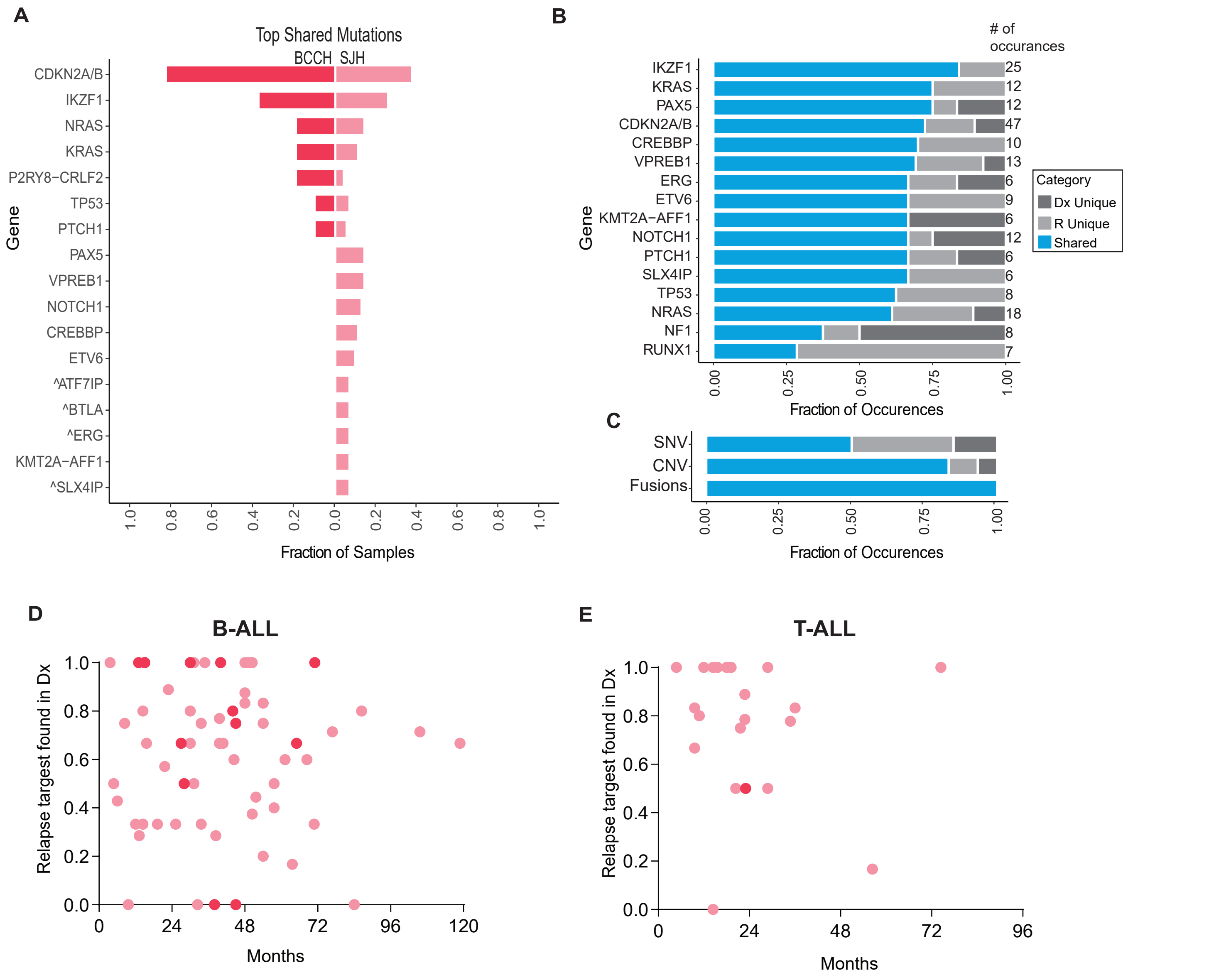


**Supplementary Figure S4: Dynamics of genomic lesions detected in the SJH and BCCH cohorts**

A. Bar plot for the most frequent variants (top 17) detected in both Dx and paired R samples (shared) plotted as the fraction of samples containing the shared variant in each cohort (n=11 from BCCH (light pink), n=69 from SJH(red)). Variants were detected in the BCCH cohort through targeted NGS while variants were detected in the SJH cohort through whole genome sequencing. Thus, variants that cannot be detected in the BCCH samples with the targeting sequencing assay are indicated with ^.

B. Bar plot for the most prevalent genes (top 16) detected in the combined BCCH and SJH cohorts. Variants are categorized as shared (blue), Dx unique (dark grey) or R unique (light grey) and plotted as the fraction of occurrences. The total number of times the gene was identified is displayed on the right end of the bar.

C. Bar plot classifying variants (SNV, CNV, Fusion) as shared (blue), Dx unique (dark grey) or R unique (light grey) and plotted as the fraction of occurrence in the BCCH cohort.

D. Dot plot for fraction of shared variants versus time to relapse for 59 B-ALL patients (n= 10 from BCCH represented by light pink circle, n= 49 from SJH represented by red circles).

E. Dot plot for fraction of shared variants versus time to relapse for 21 T-ALL patients (n= 1 from BCCH represented by light pink circle, n= 20 from SJH represented by red circles).

#### Supplementary Figure S5- BCCH Proteome Cohort- Batch Correction and Data Quality

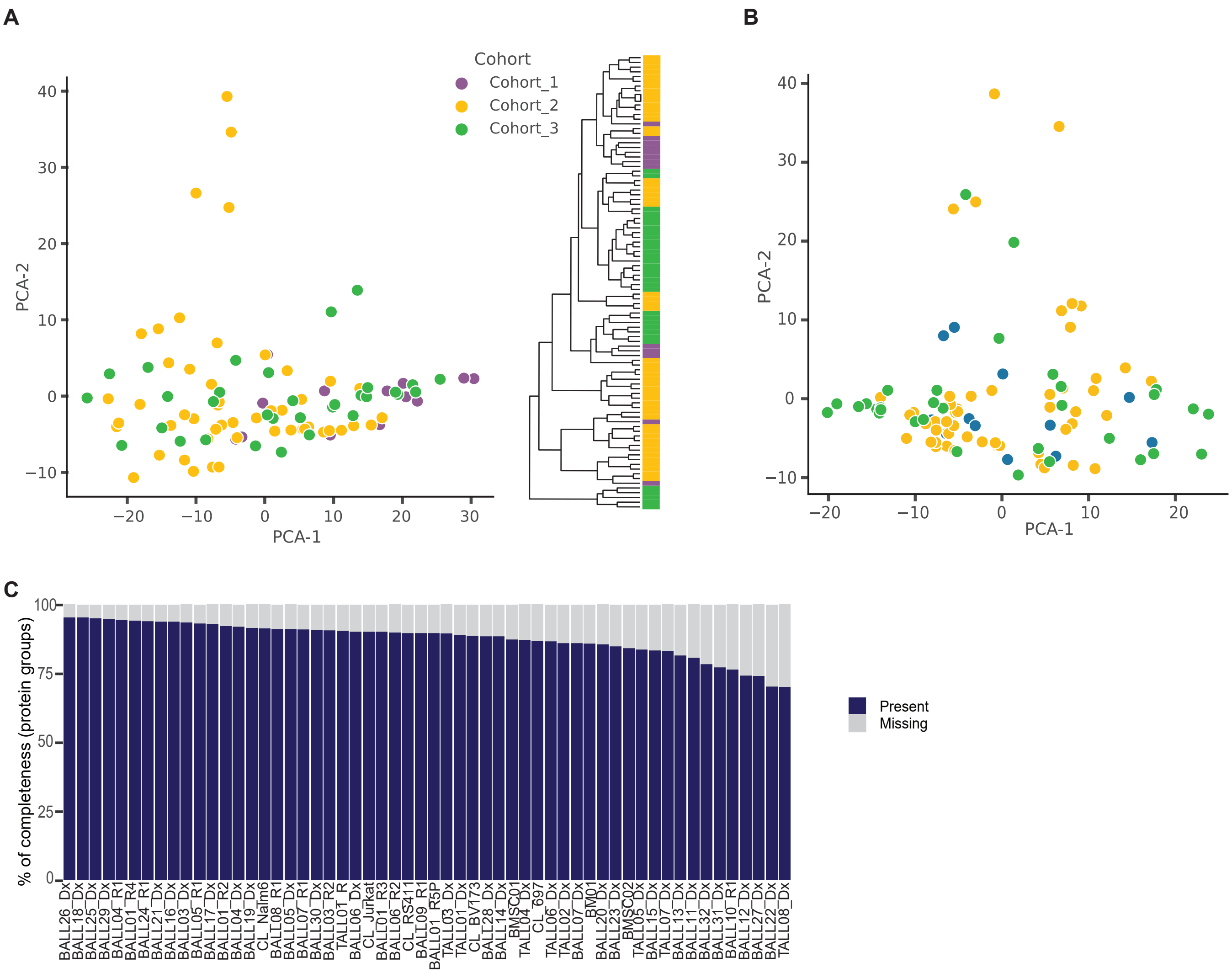


**Supplementary Figure S5- BCCH Proteome Cohort- Batch Correction and Data Quality**

**A.** (Left) PCA of proteins identified in all samples (2995) prior to batch correction. The points are colored by cohort; cohort 1 in purple, cohort 2 in yellow, and cohort 3 in green.

(Right) Hierarchical clustering of the 2995 proteins prior to batch correction scaled by min/max. The dendrogram indicates the clustering of the samples and the color bar indicates the cohort the sample was from.

**B.** (Left) PCA of proteins identified in all samples (2995) after batch correction. The points are colored by cohort; cohort 1 in purple, cohort 2 in yellow, and cohort 3 in green.

(Right) Hierarchical clustering of the 2995 proteins after batch correction scaled by min/max. The dendrogram indicates the clustering of the samples and the color bar indicates the cohort the sample was from.

**C.** Visual of data completeness in all of the samples after batch correction. Percentage of completeness is represented in blue and missing-ness is represented in grey.

**Supplementary Figure S6: Description of proteomic data filtering pipeline and quality assessment**

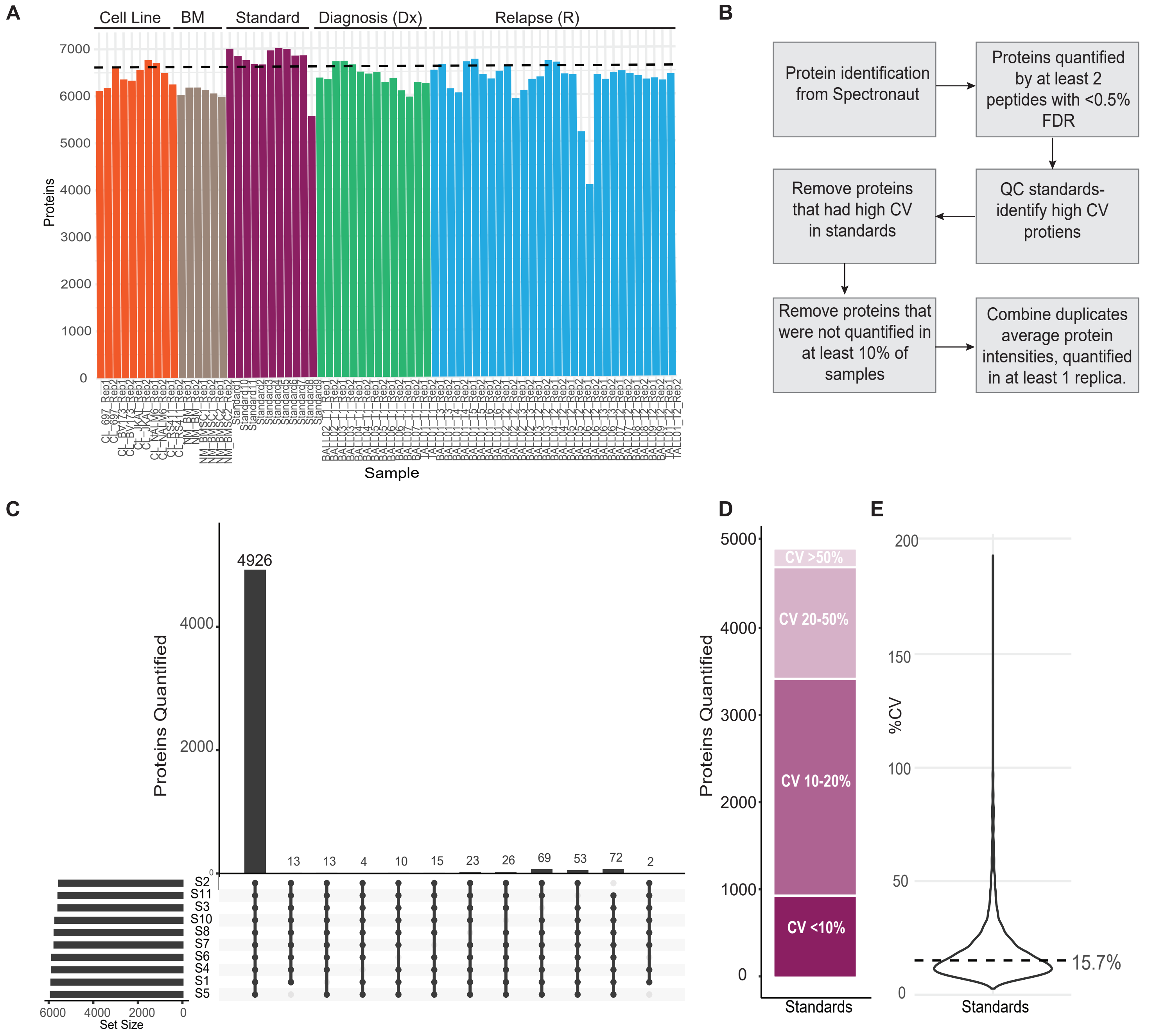


**Supplementary Figure S6: Description of proteomic data filtering pipeline and quality assessment**
**A.** Total protein groups identified in each sample, prior to any filtering. The category of the sample type is listed across the top of each group.
**B.** Diagram of the filtering workflow to attain the final proteomics dataset utilized for subsequent analyses.
**C.** Upset plot for ten of the eleven standards (standard nine was removed due to a clear technical issue). Numbers reported are based on protein groups quantified by a minimum of two peptides at a precursor q-value threshold of 0.5% FDR.
**D.** The cv for protein quantity across the ten standards was assessed and plotted by percent coefficient of variation (CV). The darkest purple bar at the bottom represents the number of proteins with a CV of less than 10% and so on, with the lightest bar at the top representing the number of proteins with a cv greater than 50%. This fraction of proteins with cv >50% was removed from the remaining data with the assumption these are unstably quantified between samples.
**E.** A violon plot demonstrating the median CV of protein quantification across the ten standards (including those >50% CV). Dashed line indicates the median CV (15.7%).

Supplementary Figure S7: Evaluation of individual sample data quality

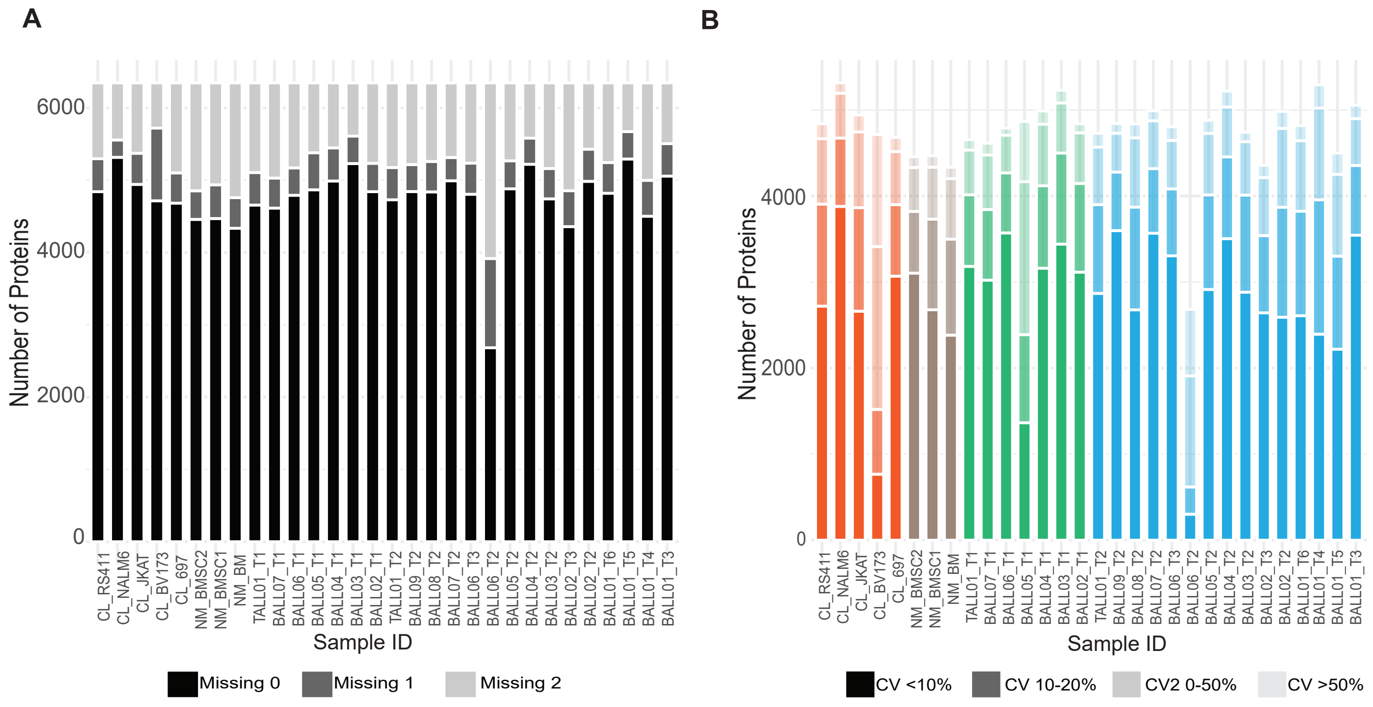


**Supplementary Figure S7: Evaluation of individual sample data quality**

A. Bar plot to demonstrate data completeness between replica of each sample. Proteins that were identified in both replica are represented in black, dark grey represents proteins that were only identified in one of the replica, and light grey represents proteins that were entirely missing from the pair.

B. CV between replica is represented as described in panel S5 D; The darkest color bar at the bottom represents the number of proteins with a CV of less than 10% and so on, with the lightest bar at the top representing the number of proteins with a cv greater than 50%.

Supplementary Figure S8: Statistical analysis of paired samples

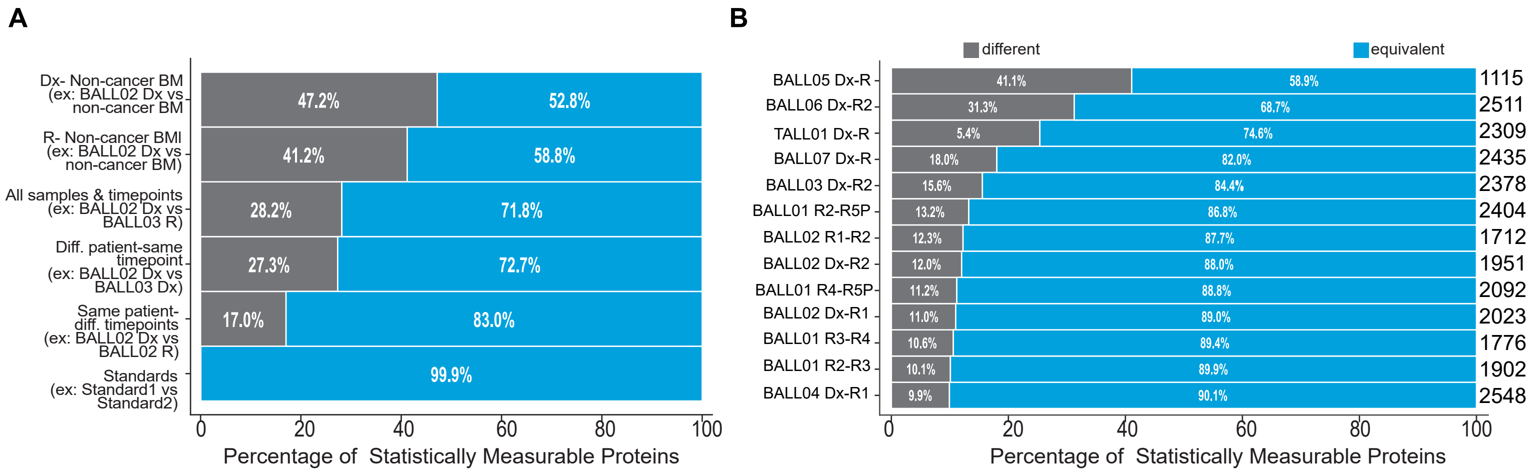


**Supplementary Figure S8: Statistical analysis of paired samples**

A. Summary of tests for differential expression and equivalence between different groups and pairings. Proteins that are statistically equivalent are represented in blue (Two-one-sided t-test (TOST) for equivalence, boundaries between log2FC<-1 and log2FC>1), proteins that are statistically different are represented in grey (student's ttest p-value<0.05, log2FC>1. The bar represents the mean equivalence or difference of all protein expression for each pairing within the group.

B. Similar representation for each individual patient pairing of the group "Same patient-diff timepoints" group. The numbers on the right of each bar indicate how many statistically measurable proteins were in each pairing.

### Supplementary Figure S9: Significant cancer associated proteins (CAPs)

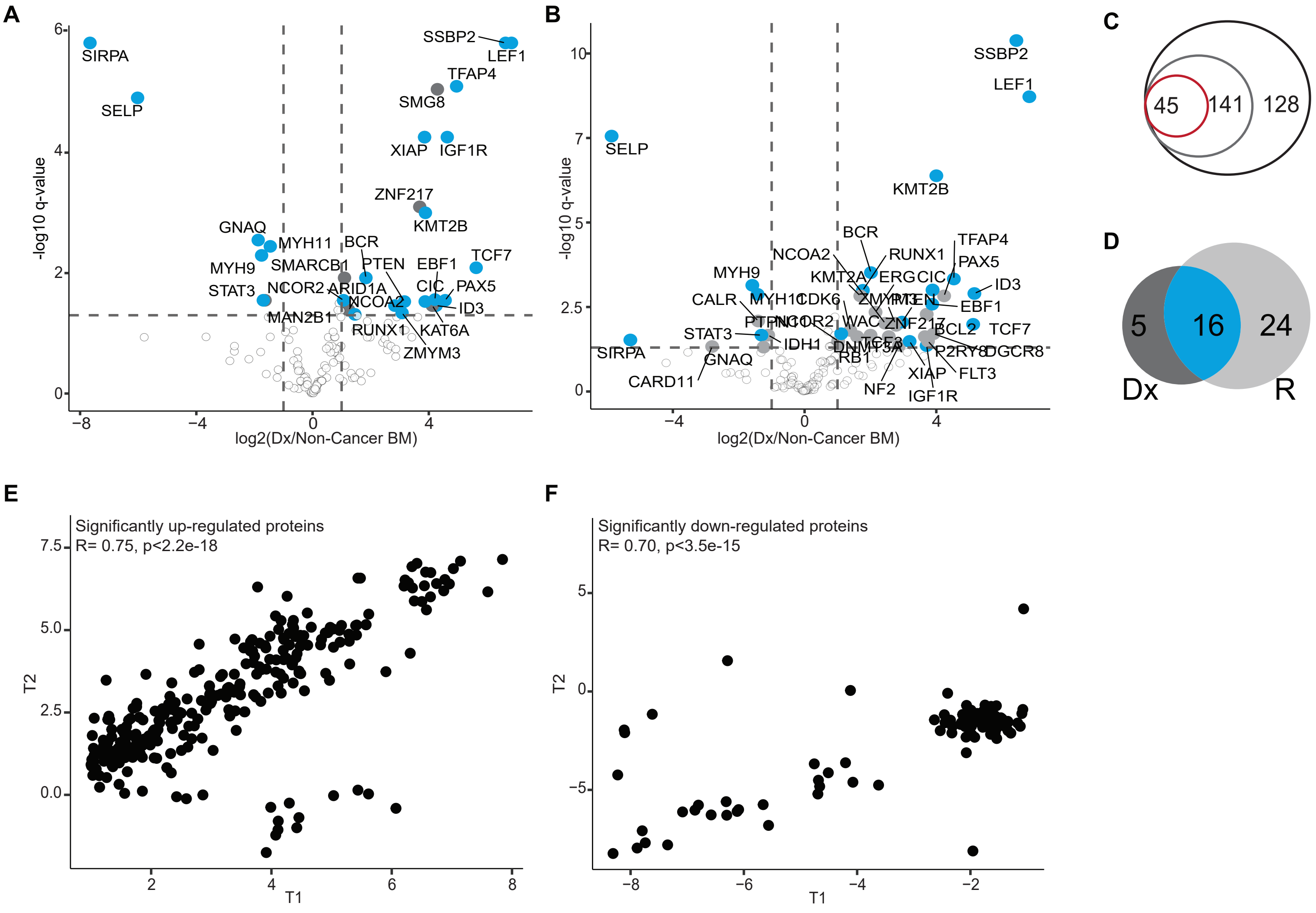


**Supplementary Figure S9: Significant cancer associated proteins (CAPs)**

A. 141 proteins were tested using LIMMA (log2FC>1, p-value adjusted FDR<0.05); Initial Diagnosis (Dx) samples vs. Non-cancer BM samples on the left and Relapse (R) vs Non-cancer BM samples on the right. Proteins that are significantly over expressed in both Dx and R are colored in blue and those that are unique to either Dx or R are colored in grey.

B. From the list of 269 pediatric cancer driving proteins (black circle), 141 proteins were detected in our data (gray circle) and 45 proteins were deemed significant (red circle).

C. Sixteen of the overexpressed proteins were shared between Dx and R (bottom)

D. Of the 45 proteins that were overexpressed at Dx, the protein expression for each protein at each timepoint was plotted as timepoint 1 (T1) vs timepoint 2 (T2) (calculated as the log2(protein expression/the average protein expression in the non-cancer bone marrow (BM) samples). A Pearson's r correlation was calculated for the entire dataset.

E. Of the 45 proteins that were under-expressed at Dx, the protein expression for each protein at each timepoint was plotted as timepoint 1 (T1) vs timepoint 2 (T2) (calculated as the log2(protein expression/the average protein expression in the non-cancer bone marrow (BM samples). A Pearson's r correlation was calculated for the entire dataset.

Supplementary Figure S10 Summary of PARP1 and yH2Ax immunofluorescence data

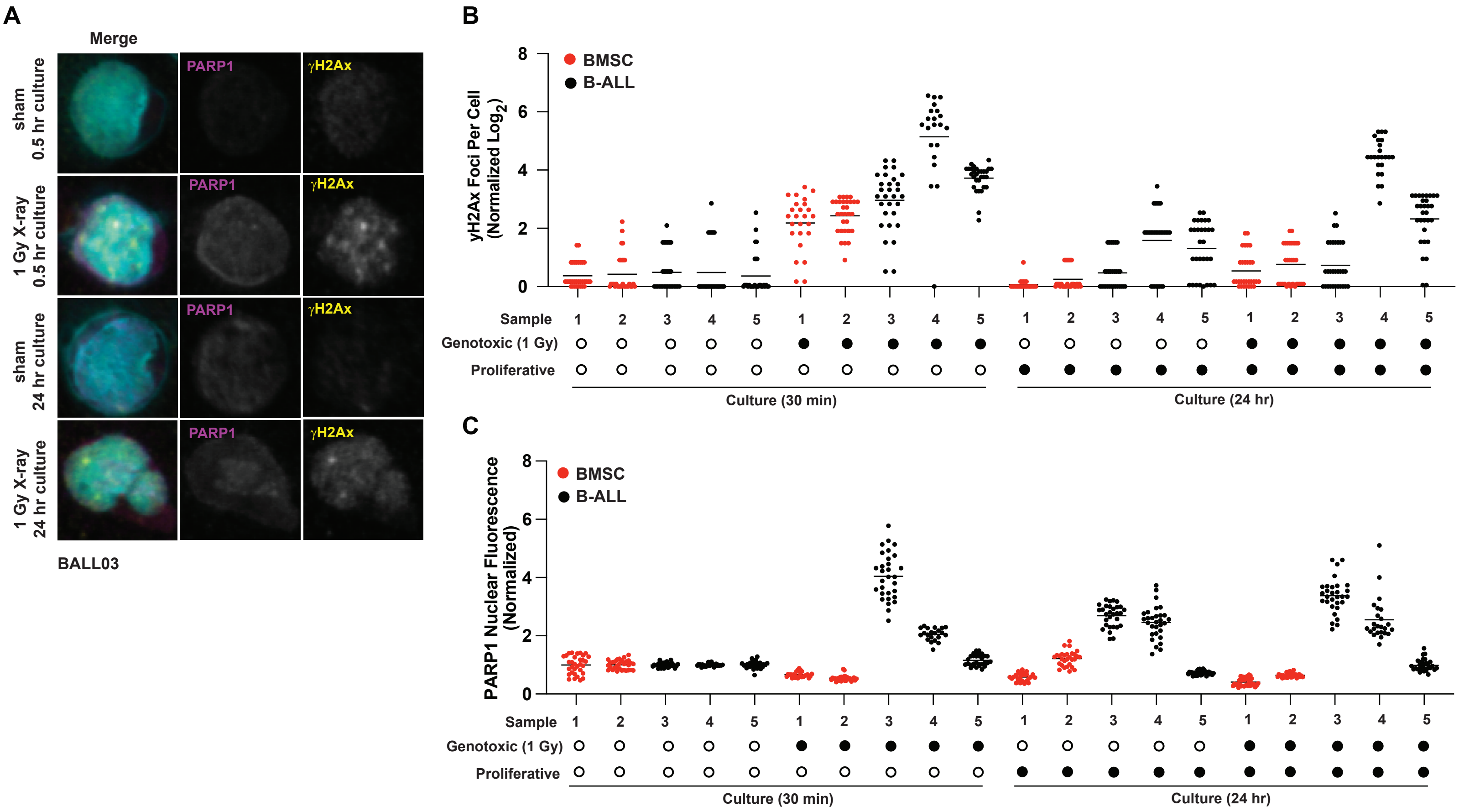


Supplementary Figure S10 Summary of PARP1 and yH2Ax immunofluorescence data

A. Representative image showing immunoflourescence staining of yH2Ax and PARP1 individually, and merged with Hoechst nuclear stain for primary sample BALL03-R2. Samples were treated with 1 Gy X-irradiation or sham conditions, and co-cultured with hTERT-MSCs for 30 minutes or 24 hours after treatment.

B. Log2 yH2Ax foci per cell normalized to sham treatment at 30 minutes, quantified from immunfluorescence analysis of 2 BMSC (red) samples and 3 B-ALL (black). Individual primary samples are indicated by number (1=BMSC02, 2=BMSC05 3= BALL04-R1, 4=BALL01-R2, 5=BALL03-R2) (n=30 cells per sample). Samples were treated with 1 Gy X-irradiation or sham conditions, and co-cultured with hTERT-MSCs for 30 minutes or 24 hours after treatment.

C. Average PARP1 nuclear fluorescence per cell normalized to sham treatment at 30 minutes, quantified from immunfluorescence analysis of 2 BMSC (red) samples and 3 B-ALL (black). Individual primary samples are indicated by number (1=BMSC02, 2=BMSC05 3= BALL04-R1, 4=BALL01-R2, 5=BALL03-R2) (n=30 cells per sample). Samples were treated with 1 Gy X-irradiation or sham conditions, and co-cultured with hTERT-MSCs for 30 minutes or 24 hours after treatment.
